# The Broad Structure of Psychopathology in the All of Us Research Program

**DOI:** 10.1101/2025.05.20.25328016

**Authors:** Alireza Ehteshami, Irwin D. Waldman

**Author notes:** **Corresponding Author:** Alireza Ehteshami, MSc, MA, Department of Psychology, Emory University, 36 Eagle Row, Atlanta, GA 30322.

## Abstract

**Background:** Given substantial comorbidity among, and considerable heterogeneity within, psychiatric diagnoses, researchers have suggested alternative systems for classifying psychopathology. The Hierarchical Taxonomy of Psychopathology (HiTOP) is a recently proposed framework for understanding mental disorders based on how symptoms and diagnoses tend to cluster across individuals. While the model is grounded in existing research and supported by recent meta-analytic evidence, its structure has not yet been directly tested using large, representative clinical datasets. In this study, we used electronic health record (EHR) data to examine the overall organization of mental disorders as proposed by HiTOP, with the goal of informing future research on biological and environmental risk factors as well as important life outcomes.

**Methods:** Data were drawn from the *All of Us* Research Program, a landmark nation-wide U.S. biobank initiative designed to advance population-scale health research, and included participants’ psychiatric diagnoses and sociodemographic correlates as documented in their EHRs. A total of 127,963 participants and 39 primary diagnoses were identified. We analyzed patterns of co-occurrence among psychiatric diagnoses to identify broader psychopathology dimensions, assess the overall structure of mental disorders, and clarify the placement of conditions that have been inconsistently categorized in past research. Several alternative dimensional models were compared based on their statistical fit, interpretability, and capacity to capture coherent and replicable patterns of comorbidity.

**Results:** A model identifying six broad and correlated dimensions—Fear, Distress, Externalizing, Substance Use, Thought Problems, and Neurodevelopmental Disorders— provided the best fit to the data. This structure was highly consistent across analyses and showed strong split-half replicability and meaningful associations with relevant clinical and demographic characteristics.

**Conclusions:** These findings support a 6-factor model of psychopathology that broadly resembles major dimensions in the HiTOP framework. By addressing key gaps in the literature, this study advances our understanding of the structure and correlates of mental disorders. The results offer a foundation for more nuanced investigations into the etiology, progression, and treatment of mental health conditions.

## Introduction

Psychiatric disorders and related traits (collectively termed psychopathology) are highly prevalent globally (Baumeister & Härter, 2007; Kessler & Wang, 2008), imposing substantial personal, interpersonal, and economic burdens (Insel, 2008). It has become evident that traditional categorical nosologies have hindered progress in gaining insight into the etiology, treatment, and prevention of psychopathology (Hyman, 2010; Kotov et al., 2017, 2021). The utility of categorical diagnoses for assessment, applied research, and clinical practice is significantly limited by factors such as pervasive comorbidity (Plana-Ripoll et al., 2019), excessive heterogeneity, low symptom specificity, poor reliability, and poor diagnostic coverage (Conway, Mansolf, & Reise, 2019; Kotov et al., 2017; Krueger et al., 2018; Wright et al., 2013). Addressing many of these limitations, the Hierarchical Taxonomy of Psychopathology (HiTOP; Kotov et al., 2017, 2021) has emerged as an alternative framework that synthesizes extant research on the structure of psychopathology. HiTOP aims to deliver reliable and valid phenotypes that can be used to advance understanding of the genetic, neurobiological, and environmental bases of psychopathology.

HiTOP organizes psychiatric signs, symptoms, and diagnoses into a dimensional hierarchy, from broad spectra to specific symptoms and traits. These hierarchical dimensions are derived by statistically analyzing how symptoms, maladaptive traits, and diagnoses co-occur across diverse samples—including clinical, community, and epidemiological populations— revealing which features consistently appear together. This analysis reveals that traditionally distinct disorders (e.g., major depression, generalized anxiety disorder, PTSD) share substantial common variance, suggesting they reflect underlying latent dimensions. The higher-order dimensions (e.g., Internalizing, Externalizing, and Thought Disorder) represent these latent constructs accounting for covariation among lower-order syndromes, symptoms, and traits. For example, the Internalizing spectrum emerges from the observed clustering of depressive, anxiety, and trauma-related disorders, hypothesized to reflect shared processes such as elevated negative affectivity, while Externalizing captures the co-occurrence of substance use disorders and antisocial behaviors, potentially reflecting common disinhibitory mechanisms.

Within this hierarchy, clinical features group into increasingly broad levels—from specific symptoms and traits to subfactors (e.g., Fear vs. Distress) to spectra—allowing the framework to explain comorbidity through shared liabilities while preserving clinically meaningful distinctions. Thus, higher-order dimensions effectively account for, and parsimoniously explain, diagnostic comorbidity by capturing common risk factors, developmental trajectories, and treatment responses (Barlow et al., 2017; Dalgleish, Black, Johnston, & Bevan, 2020; Mansell, Harvey, Watkins, & Shafran, 2009; Parkes et al., 2021), facilitating research into shared and unique etiological mechanisms while supporting transdiagnostic intervention development.

The HiTOP model was initially developed through a narrative synthesis of prior factor analytic studies, an approach that integrates findings across the literature but does not provide direct estimates of the strength or consistency of associations among disorders. Although many individual studies have supported dimensional models of psychopathology, much of this work has relied on smaller or specialized samples with limited disorder coverage—particularly for certain diagnoses such as OCD and certain neurodevelopmental disorders. These constraints limit generalizability and make it difficult to fully capture the higher-order structure of psychopathology (Kotov et al., 2017). In addition, many prior studies have modeled individual dimensions in isolation (e.g., internalizing, externalizing), rather than examining how a broader set of dimensions covary or relate to external validators such as biological correlates or treatment outcomes (Kotov et al., 2017, 2021). Most studies also have tested only a limited number of competing structural models, further restricting our ability to evaluate the relative robustness or utility of alternative dimensional frameworks (Waldman et al., 2023). A recent meta-analysis (Ringwald, Forbes, & Wright, 2023) addressed many of these limitations by quantitatively synthesizing results across dozens of studies and providing strong support for much of the HiTOP framework. Meta-analytic methods are well-suited to account for heterogeneity and increase statistical precision, but they remain limited by the available literature—particularly with respect to the range of disorders examined and the comparability of measurement models across studies. Consequently, important questions remain about the consistency and magnitude of correlations among disorders and higher-order dimensions when evaluated directly in a single, well-powered dataset. The present study addresses this need by leveraging a large, diverse sample with broad diagnostic coverage and using a comprehensive model comparison framework to test the structure and external correlates of transdiagnostic psychopathology dimensions.

To address these limitations, we utilized psychiatric diagnoses extracted from electronic health records (EHR) in the All of Us Research Program (*All of Us*)—a large and demographically diverse sample representative of the US population (Epstein, 2022; “All of Us” Research Program Investigators et al., 2019). We used Confirmatory Factor Analyses (CFAs), which test whether patterns of observed diagnostic co-occurrence fit hypothesized dimensional structures informed by HiTOP. This approach allows us to statistically evaluate the extent to which psychiatric diagnoses can be captured by interpretable higher-order dimensions, despite the fact that diagnoses themselves are heterogeneous and shaped by clinical judgment. While less granular than symptom-level data, diagnoses remain the most widely used and scalable phenotypes available in large-scale health systems, enabling investigation of broad structural patterns in real-world settings.

Within the context of evaluating the overarching HiTOP structure we addressed several specific *a priori* questions regarding the classification of psychopathology. First, we tested whether substance use disorders and non–substance-related externalizing disorders are best represented by distinct yet correlated latent dimensions. This question was motivated by dimensional models of psychopathology that conceptualize conduct disorder (CD), antisocial personality disorder (ASPD), and substance use disorders as reflecting a shared, genetically mediated liability for externalizing behavior (Krueger et al., 2021; Krueger, Markon, Patrick, Benning, & Kramer, 2007). While many latent variable studies support a unidimensional externalizing construct, others have questioned whether substance use reflects unique etiological and clinical features that warrant separate classification (Jablensky, 2009; Poore et al., 2023; Verona, Javdani, & Sprague, 2011; Voorhees et al., 2014). For instance, Verona, Javdani, & Sprague (2011) found that in a sample of adolescents, a three-factor model that distinguished Substance Use from other externalizing behaviors (e.g., conduct problems, ADHD, oppositional defiant disorder) provided superior fit compared to two-factor models, suggesting meaningful differentiation even in youth samples. Similarly, in a sample of adult veterans, Voorhees et al. (2014) identified a three-factor structure comprising Internalizing, Externalizing, and Substance Abuse, reinforcing the possibility that substance use may emerge as a separate psychopathological dimension across age groups and clinical populations. Hence, we were particularly interested in whether the inclusion of a broader and more diverse range of psychopathology indicators might allow a stronger, more meaningful distinction between substance use and other externalizing disorders to emerge—something that might not be detectable in more constrained models.

*Second*, we examined whether Distress and Fear should be represented as separate (yet related) dimensions rather than as a single overarching Internalizing dimension. This distinction is supported by research indicating that depressive and anxiety-related disorders cluster into separable, though correlated, emotional syndromes, which differ in their patterns of comorbidity, external validators, and developmental trajectories (Kotov et al., 2017; Watson, Clark, Simms, & Kotov, 2022).

*Third*, building on these specific modeling decisions, we then asked whether a model comprising six correlated higher-order factors—Fear, Distress, Externalizing, Substance Use, Thought Problems, and Neurodevelopmental Disorders—would yield superior fit relative to more parsimonious structures.

*Fourth*, we assessed whether bifactor models—often favored for their apparent superior statistical fit—would actually perform worse relative to correlated factors models on conventional model fit as well as alternative indices meant to assess the coherence and interpretability of factors. This reflects growing concern that bifactor models may capitalize on idiosyncrasies in the data and produce misleading or unstable factors (Waldman et al., 2023; Watts, Greene, Bonifay, & Fried, 2024).

*Finally*, we hypothesized that the six dimensions would be meaningfully distinct in their associations with external correlates, supporting their validity as separate constructs within a multidimensional framework of psychopathology.

In addition to testing alternative overarching structural models, we conducted targeted analyses to clarify the placement of disorders with ambiguous or debated positions within the HiTOP framework. These included examining whether ADHD better reflects Externalizing or Neurodevelopmental domains, where eating disorders and OCD fit within the hierarchy, and how personality disorders like ASPD and BPD may reflect multiple spectra. Full details of these disorder-specific analyses are provided in **eMethods in Supplement 1**.

We also refined our models to better capture relations between closely related disorders that share features beyond their primary dimensions (such as different ADHD subtypes or OCD with obsessive-compulsive personality disorder). Additionally, we used more data-driven approaches to identify potential improvements to the model, while carefully validating these changes to ensure they reflected genuine patterns rather than statistical noise (MacCallum, Roznowski, & Necowitz, 1992). These refinement procedures and validation methods are detailed in the **eMethods in Supplement 1**.

We went beyond standard statistical measures to evaluate our models in several ways. We tested whether our identified dimensions remained stable when individual diagnoses were removed, verified that our findings held true when analyzing different halves of the sample, and confirmed our models performed better than models with random arrangements of diagnoses. Finally, we examined the criterion validity of the higher-order psychopathology dimensions through their associations with relevant correlates. This project was pre-registered on the Open Science Framework (https://osf.io/y5svr/).

## Methods

### Participants

Participants for this study were drawn from the All of Us Research Program (*All of Us*), a nationwide, prospective cohort study that aims to investigate the impact of lifestyle, environment, and genomics on health outcomes. Sponsored by the US National Institutes of Health (NIH), *All of Us* aims to recruit ≥ 1 million adults over the next several years (Epstein, 2022; Investigators et al., 2019). A central focus of the program is to equitably include populations traditionally underrepresented in biomedical research (Investigators et al., 2019; Mapes et al., 2020).

Recruitment primarily occurs through participating healthcare organizations and Federally Qualified Health Centers, although interested individuals can also enroll directly at community-based sites. Participants complete necessary procedures on the All of Us website (https://joinallofus.org), including providing informed consent and completing baseline health surveys. After enrollment, participants may undergo a basic physical examination and biospecimen collection at a partnered healthcare site. Follow-up is conducted actively through periodic surveys and passively through linkage with electronic health records (EHRs).

As of October 2022, over 537,000 individuals aged 16-65 (40% male, 60% female) had joined the program. Of these participants, more than 324,000 shared electronic health records (EHRs), 372,380 completed surveys on overall health, lifestyle, and healthcare access & utilization, 311,300 provided physical measurements, and 320,000 donated at least one biospecimen. This study analyzed data from May 6, 2018, to June 6, 2022 (release 6, N = 372,380) in compliance with the All of Us Code of Conduct.

### Measures

We examined EHR lifetime psychiatric diagnoses in our primary analyses. Despite criticisms of their limitations, researchers have recently begun utilizing EHR psychiatric diagnoses in psychopathology research (Chen et al., 2018; Linder, Bastarache, Hughey, & Peterson, 2021; Smoller, 2018; Smoller et al., 2019), including in *All of Us* (Barr, Bigdeli, & Meyers, 2022). To standardize EHR data across various input sources, *All of Us* employs the Observational Medical Outcomes Partnership (OMOP) Common Data Model (Klann, Joss, Embree, & Murphy, 2019). We selected all relevant OMOP codes for psychiatric conditions (listed in **eTable 1 in Supplement 1**). Additionally, we clustered codes for highly similar diagnoses (e.g., autism spectrum disorder, autism, autistic disorder, and Asperger’s syndrome; a full list of these clusters can be found in **eTable 2 in Supplement 1**). We used sex assigned at birth, annual household income, education level, and sleep disturbance as external correlates for the psychopathology dimensions identified in our preferred structural models (**eMethods in Supplement 1** contains a full description of the external correlates).

### Statistical Analysis

Our approach involved conducting factor analyses of the EHR diagnoses on a continuum ranging from fully confirmatory to more exploratory, within a Confirmatory Factor Analytic (CFA) framework. We began by contrasting *a priori* hypothesized models to test progressive elaborations of the HiTOP structure, then tested alternative classifications of disorders with ambiguous placements, and finally used a more exploratory approach (involving modification indices) to search for cross-loadings and correlated residuals that might improve model fit. We relied primarily on correlated factor models but also contrasted these with bifactor models. In correlated factors models, indicators (i.e., diagnoses herein) load on a given factor and the resulting factors are allowed to correlate with one another. In bifactor models, all indicators load on a general factor (i.e., general psychopathology, or *p*) and on a specific factor (e.g., Internalizing or Externalizing), allowing for the separate contributions of general versus domain-specific variance in disorders. In contrast, correlated factors models assume that variance in disorders is due only to partially overlapping dimensions but not to a general factor.

Fit indices evaluate how well a factor model captures the correlational patterns in the data by quantifying the discrepancy between the covariances implied by the model and those observed in the data. Following standard practices in the field, we used multiple fit indices (CFI, TLI, RMSEA, and SRMR) to evaluate how well our dimensional models captured the structure of psychiatric disorders (see **eMethods in Supplement 1** for more details). However, accumulating evidence suggests that conventional fit indices may at times favor unnecessarily complex models or fail to detect meaningful distinctions among competing models (Bonifay & Cai, 2017; Bonifay, Lane, & Reise, 2017; Forbes et al., 2021; Greene et al., 2019; Morgan, Hodge, Wells, & Watkins, 2015; Murray & Johnson, 2013; Watts, Poore, & Waldman, 2019). To address these limitations, we supplemented conventional indices with alternative indices that evaluate three critical properties of the relationships between diagnoses and their assigned dimensions: *magnitude* (the strength of association between each disorder and its respective dimension), *precision* (the degree of statistical certainty with which these associations are estimated), and *consistency* (the extent to which the magnitudes of these associations are comparable across disorders within the same dimension) (Waldman et al., 2023). We expected these alternative indices to provide incremental utility in discriminating between competing models beyond conventional fit indices.

We examined the external validity of factors in our preferred model using simple linear regressions, estimated within structural equation models (SEMs) to characterize similarities and differences among the factors in their associations with the sociodemographic and sleep variables.

We also conducted several sensitivity analyses to assess the replicability and robustness of our preferred models. These included replicating the fit of competing models in randomly split halves of the *All of Us* sample, assessing the sensitivity of factor loadings to each factor’s constituent diagnoses, and contrasting the fit indices and alternative indices of the preferred model with models in which diagnoses were randomly assigned to the factors.

## Results

Our analytic sample comprised 127,963 individuals with at least one psychiatric diagnosis recorded in their EHRs, ensuring that all participants had been evaluated for psychopathology. The prevalence of all 39 primary diagnoses is illustrated in **eFigure 1 in Supplement 1**, and the sociodemographic information for the analytic sample is shown in **eTable 3 in Supplement 1**.

**Figure 1.**
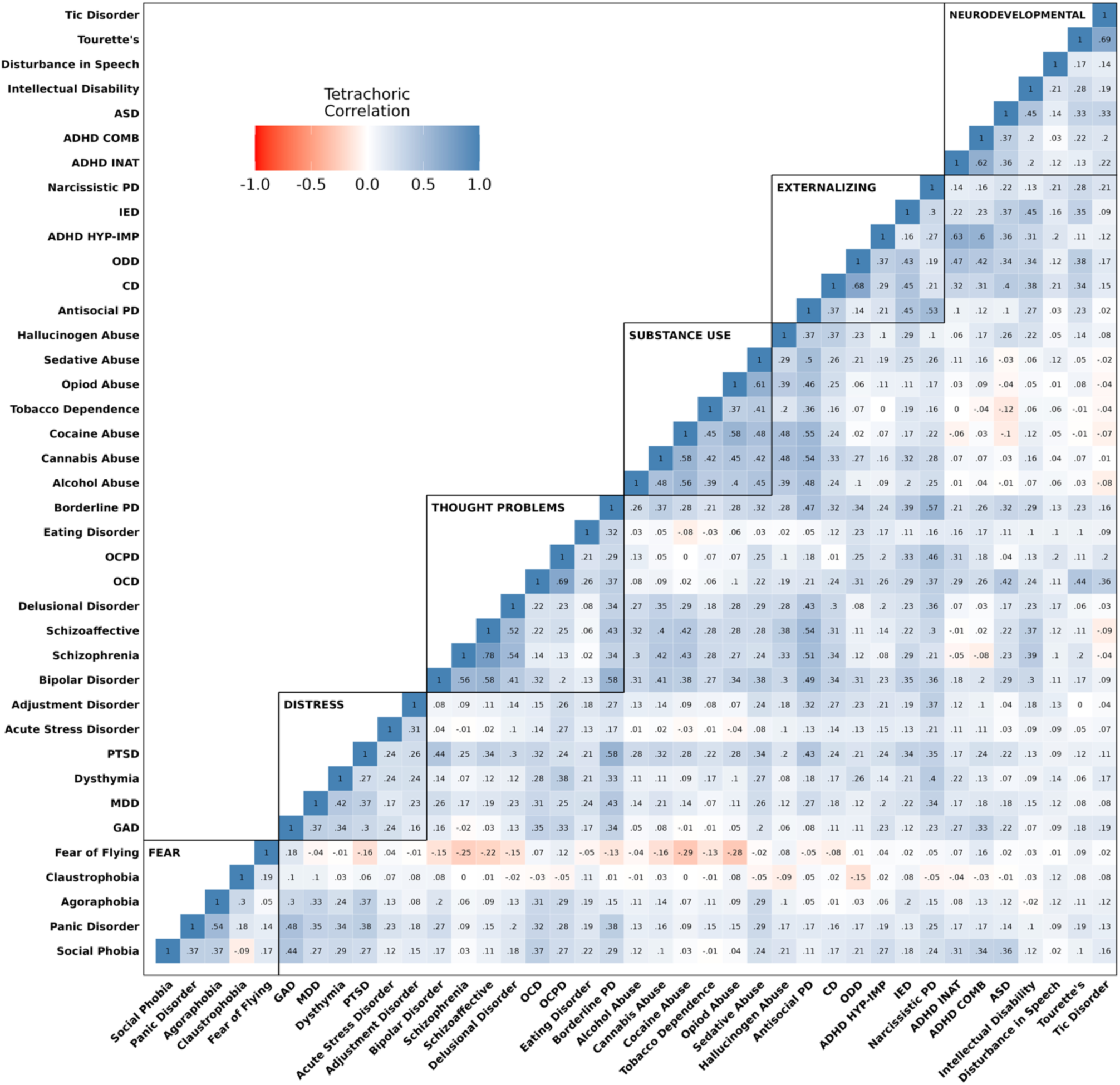
Heatmap of the correlations among 39 lifetime psychiatric diagnoses. *Note*. The heatmap is partitioned into six domains representing our most complex hypothesized model of psychopathology comprising: Fear, Distress, Thought Problems, Substance Use, Externalizing, and Neurodevelopmental Disorders.

### Correlations among Psychiatric Diagnoses

The tetrachoric correlations among the 39 psychiatric diagnoses are illustrated in the heatmap in **Figure 1**. While appreciable correlations among disorders were observed across psychopathology domains, correlations were highest among diagnoses within each domain. This pattern of correlations suggests that several correlated higher-order dimensions might best account for the comorbidity among the 39 psychiatric diagnoses.

### Tests of a priori Hypothesized Models

We next employed CFAs to test alternative hypothesized models. The main models we contrasted are displayed in **Table 1a**, and the complete set in **eTable 4 in Supplement 1** (alternative indices and factor correlations for each model can be found in **in Supplement 2**).

**Table 1.**
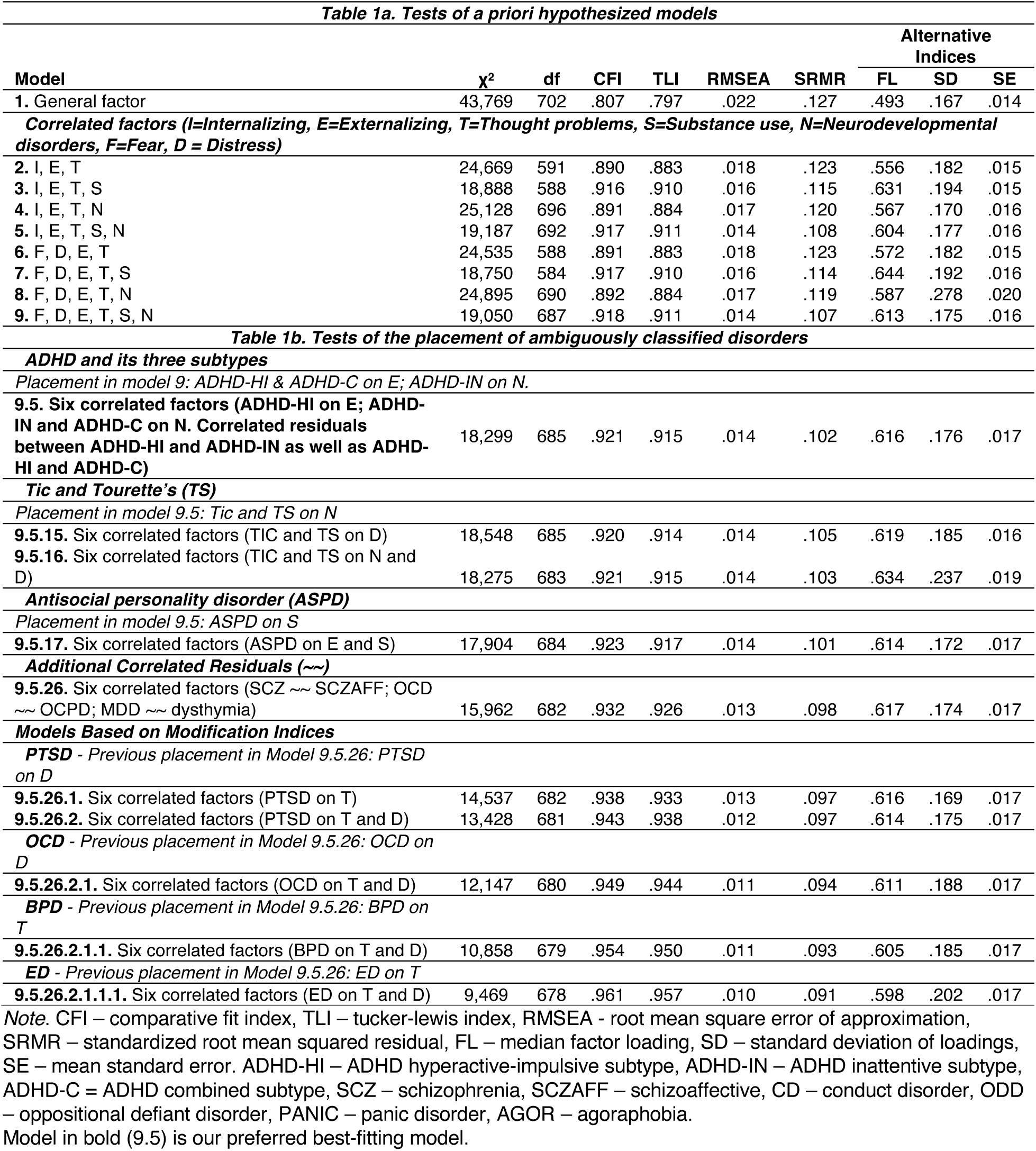
Comparison of Alternative Models of Psychopathology in All of Us.

As shown in **Table 1**, all correlated factors models (Models 2-9) demonstrated better fit than the single General factor model (Model 1). The most comprehensive model, incorporating six distinct factors (Model 9), showed the best fit. Specifically, separating Substance Use from Externalizing emerged as a critical distinction (Model 9 vs. Model 8), as reflected by both superior fit indices and alternative indices. This separation also was justified by differential associations of the external correlates with distinct Substance Use and Externalizing dimensions (discussed below), rather than an overarching combined Externalizing dimension (see **eFigure 2a in Supplement 1**). Models including a broad Internalizing dimension versus distinct Fear and Distress dimensions (Model 2-5 vs. 6-9) showed equivalent overall fit but differed in their alternative indices (shown in **eTable 5 in Supplement 1**), as discussed below.

**Figure 2.**
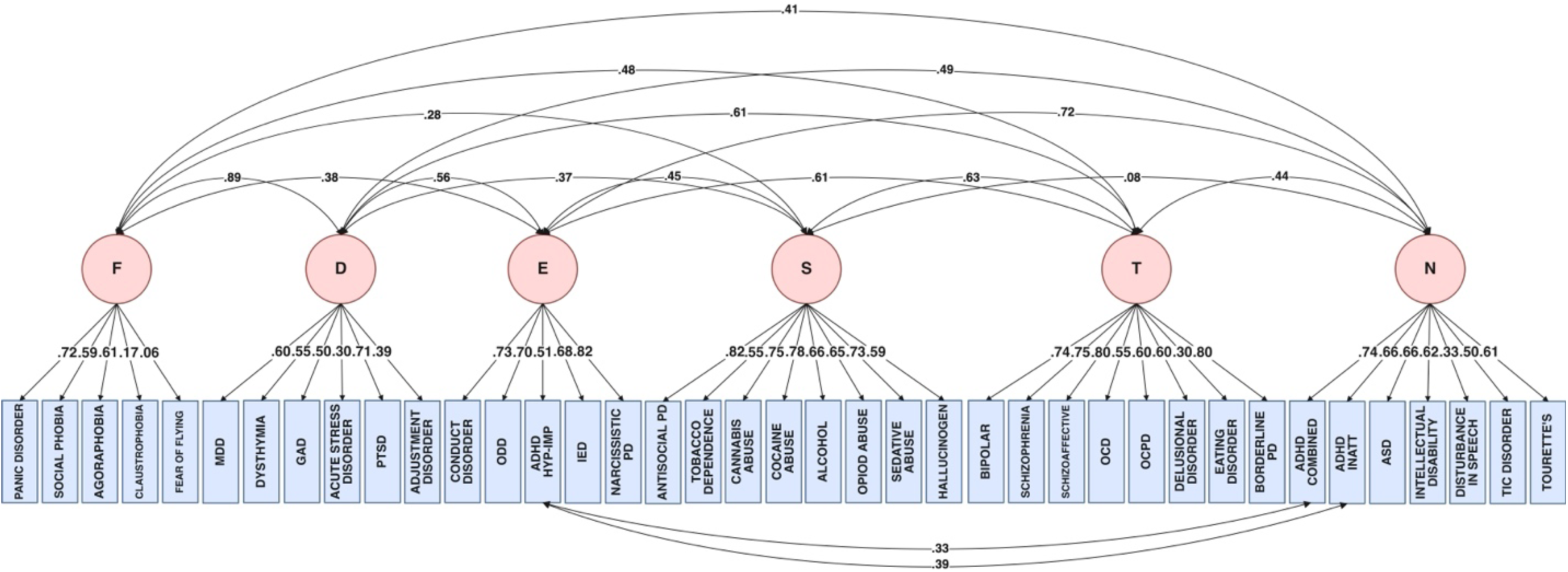
Path diagrams for preferred correlated factors model (Model 9.5). *Note*. F = Fear, D = Distress, E = Externalizing, S = Substance Use, T = Thought Problems, N = Neurodevelopmental Disorders.

Comparing correlated factors models to their bifactor equivalents revealed mixed results. While traditional fit indices generally favored bifactor models with some exceptions (**eTable 6 in Supplement 1**), alternative indices strongly supported correlated factors models, which showed more stable factor loadings with less variability and lower standard errors (for more details, see **eResults in Supplement 1**). The 6 correlated factors model (shown in **Figure 2**) performed best overall, including on traditional fit indices. These findings demonstrate the importance of using both traditional and alternative indices when evaluating structural models of psychopathology.

### Comparisons Among Factors in Their Alternative Indices

We next compared the alternative indices of the six factors from our preferred model (**Model 9.5**) to examine the extent to which the factors were well-represented by their indicators. In the preferred model, average factor loadings for diagnoses ranged from .6 to .7, with lower loadings on the Fear and Distress factors, ranging from .43 to .52. Factor loadings were estimated very precisely across all factors, with standard errors (SEs) ranging from .006 to .028. Diagnoses’ factor loadings were fairly homogeneous within factors, with standard deviations ranging from .014 for Distress to .17 for Thought Problems, except for Fear which had a standard deviation of .30. Due to the limited number of diagnoses per factor (5-8), we were unable to statistically compare these alternative indices. Detailed statistics for each factor are presented in **eTable 7 in Supplement 1**.

The distinction between Substance Use and Externalizing was supported by their alternative indices, as median Factor Loadings were higher and the variability and Standard Errors were lower in Model 9 vs. Model 8. Although models including a broad Internalizing dimension versus distinct Fear and Distress dimensions (Model 2-5 vs. 6-9) showed virtually equivalent fit, they differed in their alternative indices. As shown in **eTable 5 in Supplement 1,** for Distress diagnoses, factor loadings were comparable across the models. For Distress diagnoses, median factor loadings and standard deviations of factor loadings were similar between models. For Fear diagnoses, Model 9 showed higher median loadings with slightly increased variability. While Fear and Distress were highly correlated (r = .898), their distinct patterns of correlations with other dimensions (**eFigure 2b in Supplement 1**) support treating them as separate factors (for more details, see **eResults in Supplement 1**). The six-factor model distinguishing Substance Use from Externalizing and Fear from Distress was therefore selected for analyzing the placement of ambiguously classified disorders. Factor loadings for the diagnoses on each of the 6 factors are shown in **Table 2**.

**Table 2.**
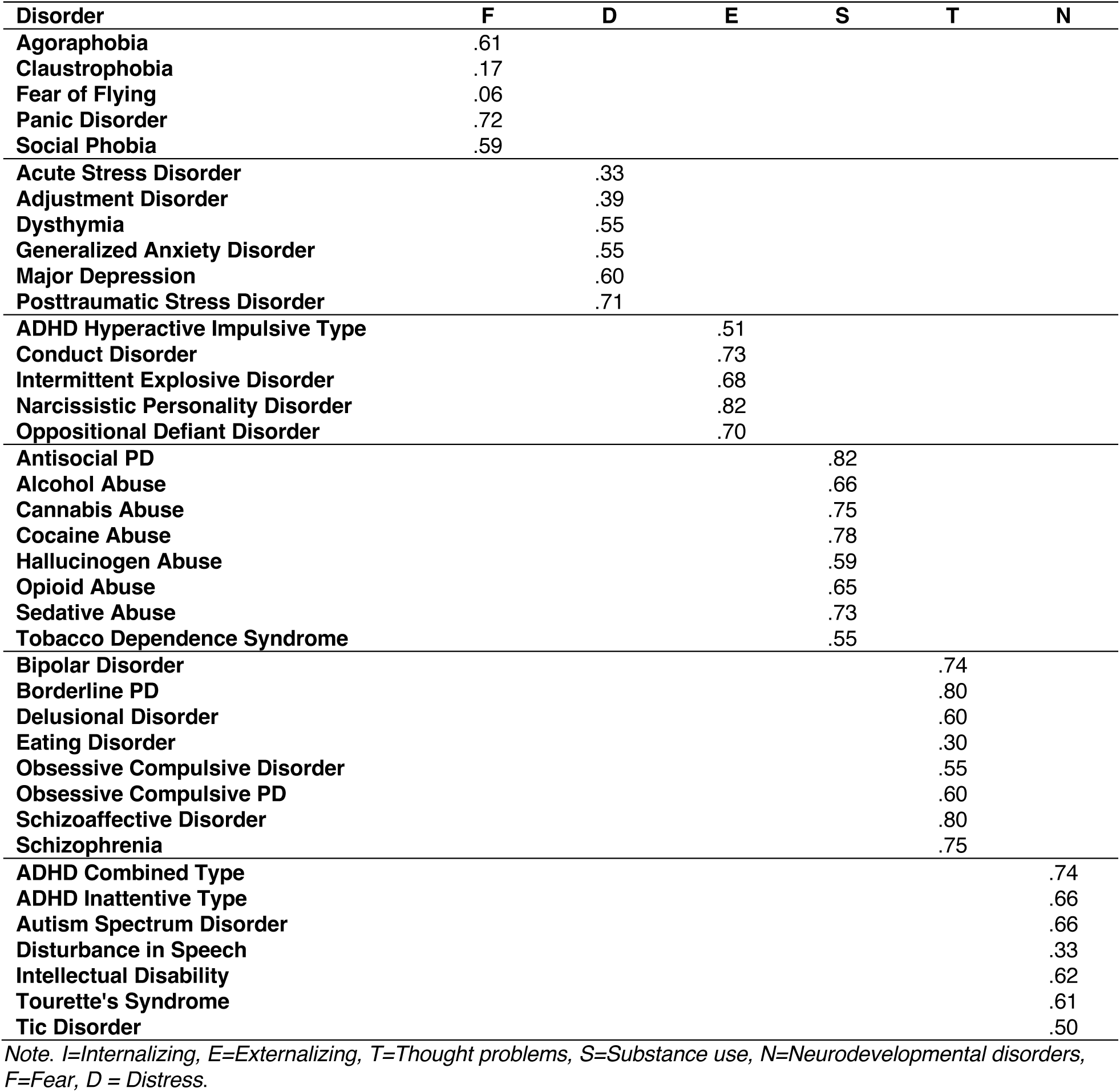
Standardized factor loadings for the preferred six-factor model of psychopathology (Model 9.5).

| Disorder | F | D | E | S | T | N |
| --- | --- | --- | --- | --- | --- | --- |
| Agoraphobia | .61 |  |  |  |  |  |
| Claustrophobia | .17 |  |  |  |  |  |
| Fear of Flying | .06 |  |  |  |  |  |
| Panic Disorder | .72 |  |  |  |  |  |
| Social Phobia | .59 |  |  |  |  |  |
| Acute Stress Disorder |  | .33 |  |  |  |  |
| Adjustment Disorder |  | .39 |  |  |  |  |
| Dysthymia |  | .55 |  |  |  |  |
| Generalized Anxiety Disorder |  | .55 |  |  |  |  |
| Major Depression |  | .60 |  |  |  |  |
| Posttraumatic Stress Disorder |  | .71 |  |  |  |  |
| ADHD Hyperactive Impulsive Type |  |  | .51 |  |  |  |
| Conduct Disorder |  |  | .73 |  |  |  |
| Intermittent Explosive Disorder |  |  | .68 |  |  |  |
| Narcissistic Personality Disorder |  |  | .82 |  |  |  |
| Oppositional Defiant Disorder |  |  | .70 |  |  |  |
| Antisocial PD |  |  |  | .82 |  |  |
| Alcohol Abuse |  |  |  | .66 |  |  |
| Cannabis Abuse |  |  |  | .75 |  |  |
| Cocaine Abuse |  |  |  | .78 |  |  |
| Hallucinogen Abuse |  |  |  | .59 |  |  |
| Opioid Abuse |  |  |  | .65 |  |  |
| Sedative Abuse |  |  |  | .73 |  |  |
| Tobacco Dependence Syndrome |  |  |  | .55 |  |  |
| Bipolar Disorder |  |  |  |  | .74 |  |
| Borderline PD |  |  |  |  | .80 |  |
| Delusional Disorder |  |  |  |  | .60 |  |
| Eating Disorder |  |  |  |  | .30 |  |
| Obsessive Compulsive Disorder |  |  |  |  | .55 |  |
| Obsessive Compulsive PD |  |  |  |  | .60 |  |
| Schizoaffective Disorder |  |  |  |  | .80 |  |
| Schizophrenia |  |  |  |  | .75 |  |
| ADHD Combined Type |  |  |  |  |  | .74 |
| ADHD Inattentive Type |  |  |  |  |  | .66 |
| Autism Spectrum Disorder |  |  |  |  |  | .66 |
| Disturbance in Speech |  |  |  |  |  | .33 |
| Intellectual Disability |  |  |  |  |  | .62 |
| Tourette's Syndrome |  |  |  |  |  | .61 |
| Tic Disorder |  |  |  |  |  | .50 |

Note. I=Internalizing, E=Externalizing, T=Thought problems, S=Substance use, N=Neurodevelopmental disorders, F=Fear, D = Distress.

### The Placement of Ambiguously Classified Disorders

We conducted targeted CFAs to determine the optimal placement of disorders with ambiguous classification (**Table 1b**; detailed procedures in **eResults in Supplement 1** and full results in **eTable 4**). For several disorders, placement decisions were informed by small differences in model fit and factor loadings. ADHD subtypes were best represented with ADHD-Hyperactive-Impulsive loading on Externalizing and both ADHD-Combined and ADHD-Inattentive loading on Neurodevelopmental Disorders, with correlated residuals between the subtypes to reflect their additional covariation not captured by the Externalizing - Neurodevelopmental Disorders factor correlation. Eating disorders showed equivalent support for loading on Distress alone or on both Distress and Thought Problems. OCD and OCPD demonstrated better fit loading on Fear or Distress rather than on Thought Problems, while Antisocial Personality Disorder better reflected Substance Use or both Substance Use and Externalizing rather than Externalizing alone. Model fit improved with Borderline Personality Disorder loading on both Distress and Thought Problems (or Externalizing) versus Thought Problems alone. Thus, these analyses were more useful in ruling out alternative models for the placement of disorders with ambiguous classification than for deciding on their optimal placement. In the final model, the addition of three theoretically-justified correlated residuals (between schizophrenia and schizoaffective disorder, OCD and OCPD, and MDD and dysthymia) to reflect their additional covariation not captured by their factor loadings improved model fit while maintaining favorable alternative fit indices.

### Replication and Robustness Across Random Sample Halves and Randomly Assigned Diagnoses

We validated our preferred 6 correlated factors model through split-half replication analyses and comparison with randomly generated alternative models. Split-half analyses demonstrated strong replication of the six-factor model results, with high intra-class correlations (.90-1.0) for most fit indices and minimal differences between halves (all < .01; **eTable 8 in Supplement 1**). Bifactor models showed much less consistency across split halves, with lower intra-class correlations for RMSEA (.79), SRMR (.85), and particularly for the mean (.56) and standard deviation (.52) of factor loadings (**eTables 9-10 in Supplement 1**). To test the specificity of diagnosis-to-factor assignments, we compared our preferred six-factor model to 1,000 random variations of the six-factor model in which diagnoses were randomly assigned to factors. Our model demonstrated superior fit across alternative indices compared to the random models (**eFigure 3 in Supplement 1**).

**Figure 3.**
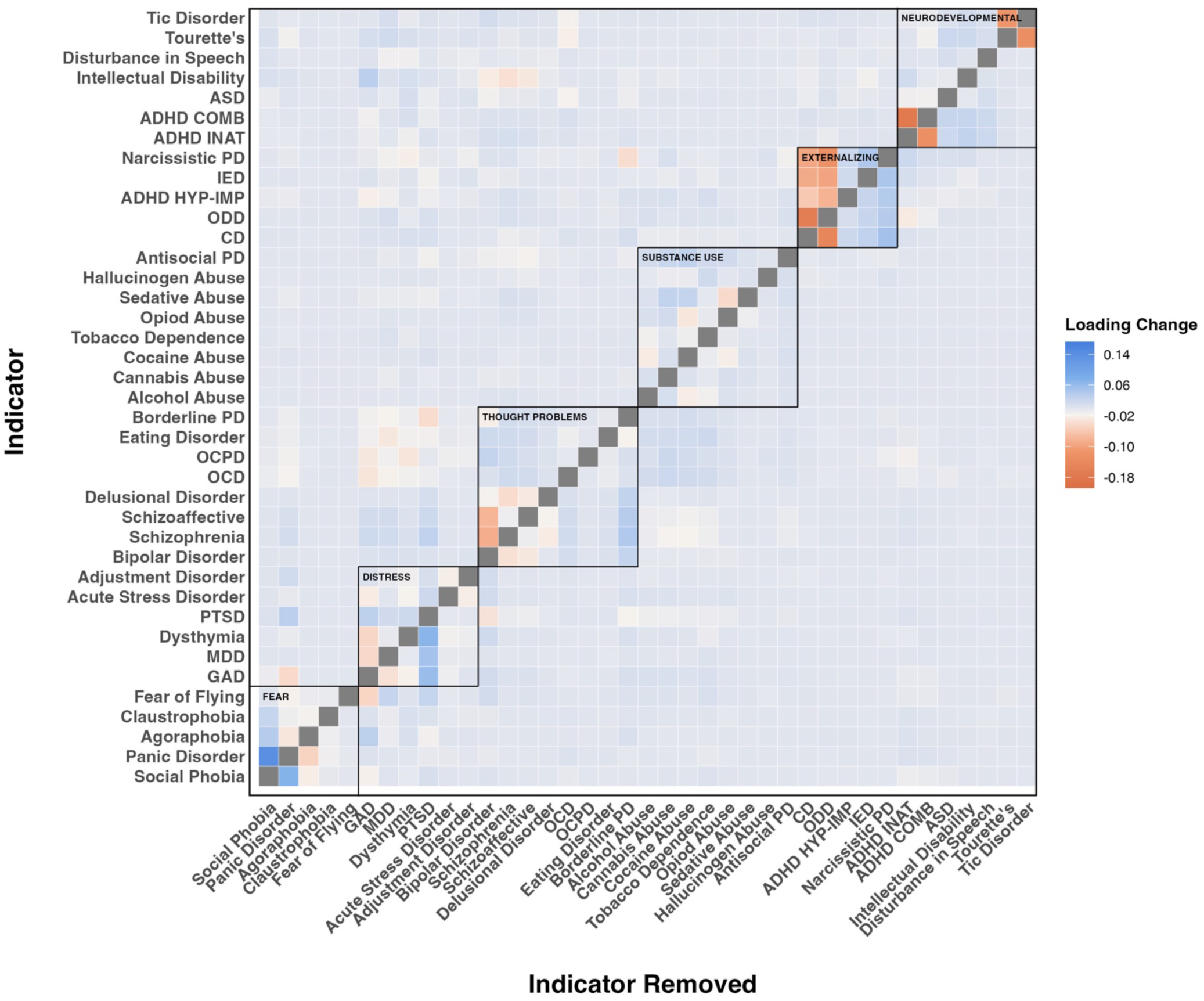
Impact of successive removal of each diagnosis on factor loadings. *Note*. Entries above the diagonal indicate the magnitude of changes in factor loadings for each diagnosis shown on the Y-axis as a function of removing each other diagnosis shown on the X-axis in turn, whereas entries below the diagonal indicate the magnitude of changes in the factor loadings for all other diagnoses shown on the Y-axis as a function of removing the diagnosis shown on the X-axis.

### The Sensitivity of Factors to Their Constituent Indicators

We examined the sensitivity of the factors to their constituent diagnoses by re-running the CFAs excluding one diagnosis at a time and examining changes in the remaining diagnoses’ factor loadings. As shown in **Figure 3**, factor loadings for diagnoses changed only slightly after removing each diagnosis in turn [M = −.0007, Median = .0002, SD = .013, minimum = −.006, maximum = .002]. Taken together, these leave-one-out CFAs indicate that the latent factors are robust: no single diagnosis exerted undue influence on the remaining loadings.

### External Correlates of the Higher-Order Psychopathology Dimensions

All associations with external correlates (i.e., sex at birth, sleep disturbance, education level, and income) significantly differed across dimensions (see **Figure 4**; all p < 5*10^-10^). Males were particularly high in Substance Use and Externalizing, whereas females were higher in Fear and Distress. Sleep disturbance showed the strongest positive association with Distress, while Substance Use showed negative associations with both education and income. Consistent with our expectations, the six psychopathology dimensions showed distinct patterns of external correlates, validating their distinction in the model. Specifically, Substance Use was more strongly associated than Externalizing with male sex and lower education and income, and the two factors showed opposite associations with sleep disturbance (positive for Externalizing, negative for Substance Use). Similarly, Distress showed stronger associations than Fear with female sex and sleep disturbance, supporting their distinction. Detailed results are presented in **eTable 11 in Supplement 1**.

**Figure 4.**
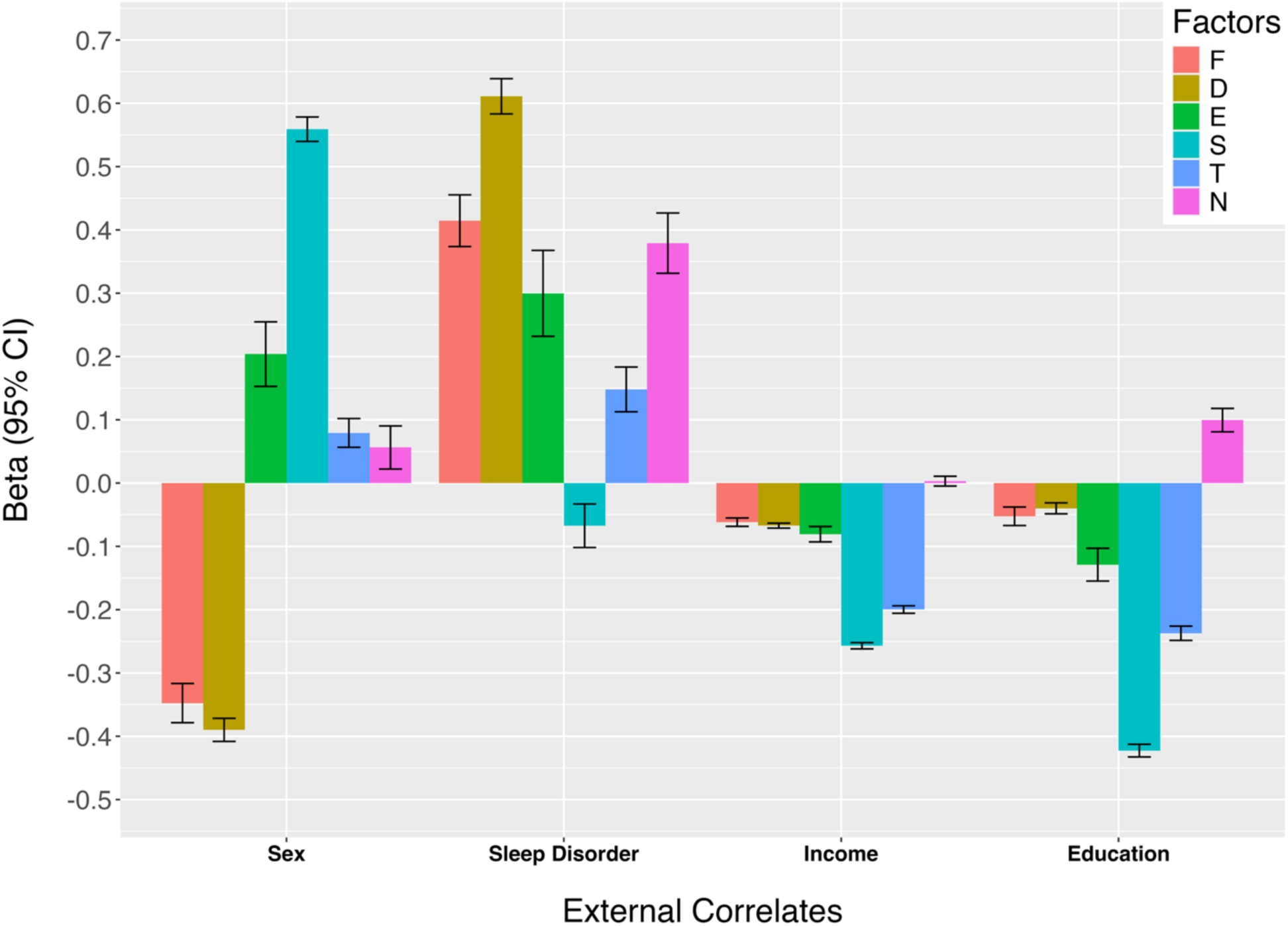
Differences in External Correlates among the 6 higher-order psychopathology dimensions (Model 9.5). *Note*. F = Fear, D = Distress, E = Externalizing, S = Substance Use, T = Thought Problems, N = Neurodevelopmental Disorders.

## Discussion

In this study, we employed both confirmatory and exploratory analyses within a CFA framework to test alternative models for the structure of psychopathology in *All of Us*. Our findings support a six-dimensional structure comprising Fear, Distress, Externalizing, Substance Use, Thought Problems, Neurodevelopmental Disorders. Correlated factors models outperformed bifactor models across multiple criteria, including replicability and fit indices. The model’s robustness was demonstrated through sensitivity analyses and replication across random sample halves. External validity analyses revealed distinct patterns of association between the six factors and demographic variables, though we did not replicate previous findings regarding educational attainment in Neurodevelopmental Disorders (Kuriyan et al., 2013; Schmengler et al., 2023; Toft et al., 2021) or sleep disturbance in Substance Use Disorders (Ara, Jacobs, Bhat, & McCall, 2016; Conroy & Arnedt, 2014; Roehrs & Roth, 2015).

Targeted CFAs suggested some refinements in the placement of OCD, OCPD, Tics, Tourette’s Syndrome, and ASPD, while ADHD subtypes showed associations with both Externalizing and Neurodevelopmental Disorders. Nonetheless, these analyses were more useful in ruling out alternative models for the placement of disorders with ambiguous classification than for deciding on their optimal placement.

The present study advances our understanding of the structure of psychopathology by addressing key gaps in the literature. Our study included a more extensive and diverse array of psychopathology indicators (i.e., diagnoses) and tested a broader range of alternative models, utilizing a sample size that surpasses nearly all extant studies in this domain. Our approach supports a hierarchical dimensional representation of psychopathology, as represented in the HiTOP model (Forbes et al., 2020; Kotov et al., 2017, 2021; Ringwald et al., 2023), addressing comorbidity and heterogeneity in categorical diagnoses using higher-order dimensions, while also representing unique variance specific to individual diagnoses. This method allows for a more nuanced characterization of psychopathology, representing both broad, overarching dimensions and specific diagnostic features, thus providing a more accurate reflection of the complex nature of mental health presentations in research and clinical practice (Conway, Forbes, et al., 2019; Conway et al., 2021; DeYoung et al., 2021).

The inclusion of a broad set of diagnoses and large sample size enabled us to test alternative placements for disorders whose classification has traditionally been ambiguous, as well as permitting more rigorous tests of bifactor models. The results of alternative indices and of split-half and random placement analyses argued against a general psychopathology factor (Fried, Greene, & Eaton, 2021; Waldman et al., 2023). Our analyses also underscored the limitations of relying solely on fit indices for model evaluation (Forbes et al., 2021; Greene et al., 2019; Waldman et al., 2023), as placing diagnoses on different factors often made very little difference to traditional fit indices, highlighting the importance of supplementing these with alternative indices. Moreover, factor correlations and external validity results supported distinguishing certain factors (e.g., Fear from Distress) even when fit indices suggested combining them (e.g., within a broader Internalizing factor). These findings demonstrate the value of a multifaceted approach to model adjudication for validly capturing the nuanced structures of psychopathology.

Despite its strengths, this study has several limitations that should be considered when interpreting the findings. **First**, while we analyzed a broad array of psychiatric diagnoses in a large, diverse sample, our reliance on diagnostic categories—rather than symptom-level data— limits the granularity of our modeling. Diagnoses are inherently heterogeneous, overlapping, and shaped by clinical conventions, which may obscure finer distinctions among syndromes.

Although recent studies using symptom-level data have generally supported higher-order structures similar to HiTOP (Forbes et al., 2020; Levin-Aspenson, 2023; Ringwald et al., 2023; Waldman & Poore, 2023), we expect that future large-scale research incorporating symptom indicators will better resolve the placement of diagnostically ambiguous conditions than we were able to herein.

**Second**, because our data were limited to DSM-based diagnoses rather than individual symptoms, we were only able to examine structure at a single level of the HiTOP hierarchy— that of broad diagnostic spectra. This prevented us from modeling the full hierarchical organization of psychopathology, including finer-grained symptom clusters or intermediate subfactors. This limitation follows directly from the nature of EHR diagnostic data and thus constrains our ability to evaluate how the structure of psychopathology may differ across levels of analysis. As future large-scale datasets begin to include rich symptom-level phenotypes, more comprehensive tests of the full HiTOP hierarchy will become possible.

**Third**, our modeling approach imposed certain constraints that warrant consideration. For example, in line with traditional confirmatory factor analysis, we initially prohibited cross-loadings of diagnoses on multiple factors. Although a limited number of theory-driven cross-loadings were introduced during model refinement, this constraint may have artificially increased the number of latent dimensions needed to account for comorbidity among diagnoses. More flexible methods—such as exploratory structural equation modeling (ESEM)—may offer a different representation of the underlying structure, particularly for diagnostically complex conditions that span multiple domains.

**Fourth**, while our preferred model demonstrated strong overall fit and robustness, several alternative models yielded similarly acceptable fit using conventional indices.

Consequently, decisions regarding the placement of certain disorders—particularly those with ambiguous nosological status—were partially guided by theoretical plausibility amid small empirical differences. This underscores the fact that fit indices may be particularly useful for winnowing out less viable models than for selecting a “best” model (Waldman et al., 2023) and highlights the interpretive nature of structural model adjudication and reinforces the need for replication and triangulation across methods.

**Fifth**, despite its importance, we did not formally evaluate whether the factor structure varied across demographic subgroups using measurement and structural invariance analyses. Although we examined associations between latent dimensions and variables such as sex, income, and education, we did not assess how such associations may vary across demographic groups. Given the demographic diversity of the *All of Us* cohort, future research should test whether the structure of psychopathology generalizes across sex, socioeconomic strata, and racial or ethnic groups.

**Sixth**, our use of diagnoses extracted from EHRs also presents several unresolved challenges, including concerns about reliability, validity, and the optimal algorithms for diagnostic classification (Chen et al., 2018; Davis, Sudlow, & Hotopf, 2016; Denny et al., 2013; Hripcsak & Albers, 2012; Linder et al., 2021; Smoller, 2018; Wei et al., 2017, 2016; Wu et al., 2019; Zheutlin et al., 2019). Diagnostic practices vary across providers, health systems, and time, and are influenced by differences in access to care, comorbidity documentation, and clinician judgment. These sources of variability may affect the completeness and consistency of diagnosis-based phenotyping. Despite these limitations, EHR-based diagnoses serve a valuable practical purpose by enabling scalable access to clinically meaningful psychiatric data in very large and diverse samples. They offer a foundation for constructing more representative cohorts and open the door to powerful downstream opportunities—such as integrating hierarchical dimensional models like HiTOP with the extensive genetic, biological, behavioral, and environmental data available in large-scale biobanks such as *All of Us*. As methodological innovations—including natural language processing, machine learning, and probabilistic phenotyping—continue to improve the validity of EHR data (An et al., 2023; Dahl et al., 2023; Papini et al., 2023; Weng et al., 2024; Yun et al., 2024), their utility in psychiatric research is likely to be greatly enhanced.

**Finally**, the structure of psychopathology represented in our preferred model may be influenced in part by our reliance on lifetime diagnoses. While this approach captures a broad history of disorder, it fails to account for the time-specific manifestations and fluctuations of symptoms across the lifespan. Psychopathology is known to exhibit age-related shifts in presentation, severity, and comorbidity patterns, particularly during key developmental transitions such as adolescence and late adulthood (Rutter, 2013; Skodol & Bender, 2016). As such, lifetime diagnoses may obscure important temporal dynamics, including periods of remission, recurrence, or the emergence of new symptoms/syndromes. Future research should aim to incorporate longitudinal diagnostic data to model within-person changes over time, ideally using large, developmentally diverse samples. Leveraging repeated measures, time-varying covariates, and growth modeling techniques could help clarify how the structure and correlates of psychopathology evolve, and the extent to which transdiagnostic dimensions remain stable or shift across developmental stages. Such approaches are important not only for refining nosological models like HiTOP but also for informing prevention and intervention strategies tailored to individuals at different developmental stages.

Hierarchical dimensional models, such as HiTOP, offer a framework that parsimoniously represents patterns of comorbidity at higher levels of the taxonomy while preserving critical distinctions at lower levels. Unlike traditional diagnostic systems like the DSM, which collapse individual differences into broad, heterogenous, highly comorbid categories, this hierarchical dimensional approach explicitly maintains individual differences at multiple levels, enabling analyses of psychopathology at different levels of generality and specificity. The comprehensive model we propose herein leverages these advantages to provide a foundation for more nuanced investigations into the causes and outcomes of both broad, higher-order psychopathology dimensions and more specific, lower-order factors. This multi-tiered approach is particularly valuable for examining similarities and differences in psychopathology across diverse ethnicities (Cicero & Ruggero, 2021; Eaton et al., 2013; Forbes et al., 2021; He & Li, 2021; He, Rodriguez-Seijas, Waldman, & Li, 2023; Moriarity, Joyner, Slavich, & Alloy, 2022; Ringwald, Forbes, & Wright, 2022; Rodriguez-Seijas et al., 2023), as well as exploring how genetic and environmental risk factors may influence psychopathology at various levels of the hierarchy across different ancestral backgrounds.

## Data Availability

All of Us data are available to researchers via application at the following link: https://www.researchallofus.org/register/

## Acknowledgements

We gratefully acknowledge *All of Us* participants for their contributions, without whom this research would not have been possible. We also thank the National Institutes of Health’s *All of Us* Research Program for making available the participant data used in this study.

## Author Contribution

All authors contributed to and have approved the final manuscript.

