## Supplement 1 for "The Broad Structure of Psychopathology in the All of Us Research Program"

### Supplemental Online Content


| | |
|---|---|
| **eMethods** | **p.2** |
| **eResults** | **p.3** |
| **eReferences** | **p.6** |
| **eTable 1.** Names and concept codes for initial set of selected diagnoses | **p.9** |
| **eTable 2.** Frequencies and diagnostic codes for psychiatric conditions used in analyses | **p.12** |
| **eTable 3.** All of Us sample characteristics across race-ethnicity | **p.13** |
| **eTable 4.** Fit and alternative indices for confirmatory and exploratory models | **p.15** |
| **eTable 5.** Alternative indices for fear and distress diagnoses across confirmatory models | **p.17** |
| **eTable 6.** Fit and alternative indices for bifactor and modified bifactor versions of best fitting models | **p.18** |
| **eTable 7.** Comparisons among factors in Model 9.5 in their alternative indices | **p.19** |
| **eTable 8.** Split-half replication of all models | **p.20** |
| **eTable 9.** Split-half replication of exploratory bifactor models | **p.22** |
| **eTable 10.** Split-half replication of exploratory modified bifactor models | **p.23** |
| **eTable 11.** External correlates of the higher-order psychopathology dimensions | **p.24** |
| **eFigure 1.** Prevalences of the 39 primary lifetime diagnoses | **p.25** |
| **eFigure 2a.** Correlations of D, F, N, and T factors with Distinct Externalizing, Substance Use, and Combined Externalizing + Substance Use factors | **p.26** |
| **eFigure 2b.** Correlations of E, N, S, and T factors with Fear, Distress, and a broad Internalizing factor | **p.26** |
| **eFigure 3.** Indices of robustness of the results of the best-fitting model using random models | **p.27** |

#### eMethods

##### *Sociodemographic correlates*

- Sex assigned at birth: Female, Male
- Annual household income: under $10k, $10k-$25k, $25k-$35k, $35k-$50k, $50k-$75k, $75k-$100k, $100k-$150k, $150k-$200k, more than $200k
- Educational attainment: Less than a high school degree or equivalent; Highest Grade: Twelve or GED; Highest Grade: College One to Three; College graduate or advanced degree

##### *Targeted analyses for ambiguously classified disorders*

In addition to these overarching structural hypotheses, we conducted several targeted CFAs and exploratory analyses to address the ambiguous placement of specific disorders within the existing psychopathology literature. These analyses were designed to clarify how certain disorders align with, or deviate from, broader latent dimensions. First, we tested a priori hypothesized correlated residuals among ADHD subtypes to account for any covariation not fully explained by their shared factor loadings. Second, we examined whether ADHD and its subtypes are best conceptualized as part of the Externalizing spectrum, the Neurodevelopmental domain, or both. Third, we investigated whether Eating Disorders reflect underlying Internalizing pathology (particularly Distress) and/or Thought Problems. Fourth, we assessed whether Obsessive-Compulsive Disorder (OCD) and Obsessive-Compulsive Personality Disorder (OCPD) load on Neurodevelopmental Disorders, Internalizing, Fear, Distress, and/or Thought Problems, reflecting ongoing debate about their nosological placement. Fifth, we evaluated whether Tics and Tourette's Syndrome align more closely with Neurodevelopmental Disorders or Distress. Sixth, we tested whether Antisocial Personality Disorder (ASPD) is best represented by Externalizing, Substance Use, or both (Verona, Javdani, & Sprague, 2011; Voorhees et al., 2014). Seventh, we explored whether Borderline Personality Disorder (BPD) reflects Distress, Thought Problems, and/or Externalizing, given its complex and multifaceted clinical presentation.

To further evaluate potential model misspecifications, we introduced theoretically and empirically justified correlated residuals between closely related disorders, including ADHD subtypes, OCD with OCPD, Schizophrenia with Schizoaffective Disorder, and Major Depressive Disorder (MDD) with Dysthymia. Finally, in an empirically driven effort to refine model fit, we used Modification Indices (MIs)—which quantify expected model improvement based on the change in $c^2$—to guide the addition of correlated residuals and cross-loadings. Importantly, cross-loadings were added only when there was clear theoretical and/or empirical support in the existing literature. Given the known risks of overfitting and capitalization on chance inherent in MI-guided modifications (MacCallum, Roznowski, & Necowitz, 1992), we evaluated all added parameters using alternative fit indices and tested their robustness through split-half replication procedures.

##### *Statistical analyses*

Our approach involved conducting factor analyses on a continuum ranging from fully confirmatory to more exploratory, using Confirmatory Factor Analyses of the binary diagnoses. We employed the Weighted Least Squares Means and Variance Adjusted Satterthwaite estimator (WLSMVS)(Flora & Curran, 2004; Muthén, 1984; Shi, DiStefano, McDaniel, & Jiang,

2018) in the R (4.4.0) package *lavaan* (0.6-17)(Loehlin & Beaujean, 2017; Rosseel, 2012) on the *All of Us* Workbench to test and replicate more comprehensive models of psychopathology than was previously possible.

Our tested models represent a substantial subset of the Hierarchical Taxonomy of Psychopathology (HiTOP)(Conway, Forbes, South, & Consortium, 2021; DeYoung et al., 2021; Kotov et al., 2017, 2021; Krueger et al., 2021; Ringwald, Forbes, & Wright, 2023), an overarching framework supported by extensive research. We tested several major components of this model, focusing on the higher-order dimensions of Externalizing (E), Internalizing (I), Distress (D), Fear (F), Substance Use Disorders (S), Thought Problems (T), and Neurodevelopmental Disorders (N). In addition to contrasting *a priori* models, we used modification indices to search for cross-loadings that might enhance model fit.

To assess model fit, we employed multiple indices given their desirable properties: the Comparative Fit Index (CFI)(Bentler, 1990), Tucker-Lewis Index (TLI)(Tucker & Lewis, 1973), Root Mean Square Error of Approximation (RMSEA) with its 95% confidence interval(Browne & Cudeck, 1992), and Standardized Root Mean Squared Residual (SRMR)(Bentler, 1995; Loehlin & Beaujean, 2017). Standard guidelines representing adequate and good fit for these fit indices are CFI and TLI values > .90 and > .95, respectively; RMSEA values of < .08 and < .05, respectively; and SRMR < .08(Li-tze Hu & Bentler, 1998; Li-tze Hu & Bentler, 1999). Researchers have recently documented the limitations of conventional fit indices (e.g., overfitting, differential fit propensity, and bias in tests of certain models) for adjudicating among structural models of psychopathology and have suggested alternative criteria(Bonifay & Cai, 2017; Bonifay, Lane, & Reise, 2017; Forbes et al., 2021; Greene et al., 2019; Morgan, Hodge, Wells, & Watkins, 2015; Murray & Johnson, 2013; Watts, Poore, & Waldman, 2019). Hence, we augmented these fit indices with additional indices for adjudicating among alternative models: the mean, median, standard deviation (SD), and standard errors (SE) of standardized factor loadings, as well as the sensitivity of factor loadings to each factor's constituent diagnoses. Superior models are characterized by higher mean and median factor loadings with lower SEs and SDs, as well as factor loadings that are less sensitive to the removal of any single diagnosis(Waldman et al., 2022). We anticipated that these alternative indices would provide incremental utility beyond conventional fit indices in discriminating between competing models.

#### eResults

##### *Distinguishing Fear and Distress within Internalizing*

Several models showed similar overall fit, particularly the model treating Internalizing as a single dimension compared to the model splitting it into Fear and Distress (Hypothesis 2: Models 5 vs. 9). Both models had identical fit indices but differed in alternative indices for relevant diagnoses (shown in eTable 5 in Supplement 1). For Distress-related diagnoses, median and standard deviations of factor loadings were similar between models (.545 and .121 for Model 5 vs. .547 and .140 for Model 9), though Model 5 had lower standard errors (.015 vs. .007). For Fear-related diagnoses, Model 9 demonstrated higher median factor loadings (.591 vs. .535) with slightly greater standard deviations (.298 vs. .268) and standard errors (.022 vs. .020). These results underscore the benefit of distinguishing between Fear and Distress to better capture diagnostic variability. Additionally, as shown in eFigure 2b in Supplement 1, correlations of Externalizing, Substance Use, Thought Problems, and

Neurodevelopmental Disorders with broad Internalizing and Distress were similar, but correlations with Fear were notably lower. Despite a high correlation between Fear and Distress (r = .898, SE = .011), these results indicate that Fear and Distress are distinct dimensions. Consequently, the six-factor model distinguishing these dimensions was identified as the preferred framework for analyzing ambiguously classified disorders.

##### *Correlated factors models vs. bifactor models*

A mixed picture emerged when comparing the correlated factors models to their bifactor model equivalents that also contained a general factor (shown in eTable 6). While conventional fit indices favored most bifactor models (given higher CFI and TLI, equivalent RMSEA, and lower SRMR), the alternative indices strongly favored the correlated factors models. Specifically, the correlated factors models demonstrated higher median factor loadings and lower standard deviations and standard errors, and these indices were much less consistent across random halves in the bifactor models. Notably, the 6 correlated factors model outperformed the corresponding bifactor model even according to conventional fit indices. These results further support the 6 correlated factors model (shown in Figure 2) as the best-fitting a priori hypothesized model. Moreover, these findings highlight the utility of alternative indices and factor correlations in augmenting conventional fit indices for adjudicating among alternative structural models of psychopathology.

##### *The Placement of Ambiguously Classified Disorders*

We examined the fit and alternative indices of several targeted CFAs to resolve the placement of ambiguously classified disorders (Table 1 in the main text shows results for disorders whose placement could be resolved, whereas eTable 4 in Supplement 1 shows the full set of results).

Differences in fit for the placement of ADHD or its subtypes were small and thus hard to adjudicate (Models 9.1 - 9.6). Having ADHD load on both E and N or having ADHD-Hyperactive-Impulsive subtype (ADHD-HI) load on E and ADHD-Combined subtype (ADHD-C) and ADHD-Inattentive subtype (ADHD-IN) load on N with correlated residuals between ADHD-HI and the other two subtypes, showed similar fit. The latter model (Model 9.5) was preferred, given that the loadings of the ADHD subtypes were substantially higher, commensurate with the other diagnoses' loadings on E and N.

The placement of eating disorders (including Anorexia Nervosa and Bulimia Nervosa; Models 9.5.1 - 9.5.5) could not be definitively resolved, as both model fit and alternative indices similarly favored their placement on either D only, or D and T. There was nearly equivalent support for a model in which anorexia loaded on D and bulimia loaded on D and E.

Despite similarities in model fit and alternative indices, there was some suggestion that OCD and OCPD might reflect F or D, rather than T (Models 9.5.6 - 9.5.9). Results for Tics and Tourette's Syndrome (Models 9.5.15 and 9.5.16) suggested they may be better placed on D instead of (or in addition to) N.

Antisocial Personality Disorder appeared to better reflect S, or S and E, rather than E only, although differences in factor loadings and correlations for these models were minimal (Models 9.5 and 9.5.17). Borderline Personality Disorder appeared to reflect D in addition to T or E, rather than T alone (Models 9.5.10 - 9.5.14).

Based on the high similarity of their diagnoses, eight correlated residuals were considered *a priori* for addition to the preferred 6 correlated factor model (Models 9.5.18 - 9.5.26). When added simultaneously to Model 9.5, three correlated residuals (between schizophrenia and schizoaffective disorder, OCD and OCPD, and MDD and dysthymia) improved model fit while maintaining equivalent levels of the alternative indices. Finally, Modification Indices increased conventional fit indices only slightly while worsening alternative indices.

##### *Replication and Robustness Across Random Sample Halves and Randomly Assigned Diagnoses*

We tested for replication of the model fitting results across random halves of the *All of Us* sample. As shown in eTable 8, findings were highly similar across the random halves and mirrored those from the full sample. Models demonstrated better fit and more favorable alternative indices with increasing model complexity. Specifically, intra-class correlations of the fit and alternative indices across random halves of the sample were very high (.90 to 1.0), except for mean factor loadings (FLs) which had a correlation of .765. Differences across random halves in the indices were minimal, ranging from .009 for FL standard deviations (SDs) to .0026 for the Tucker-Lewis Index (TLI). These indices were much less consistent for the bifactor models across random halves (eTable 9 and eTable 10). Although intra-class correlations of the CFI and TLI were similarly high (.93), these correlations were lower for RMSEA and SRMR (.79 and .85) and for the mean and SD of factor loadings (.56 and .52). Differences in CFI, TLI, RMSEA, and SRMR were minimal (.00009 to .0023), but greater for the mean and SD of factor loadings (.024 and .048).

To address concerns that our preferred six-factor models might have the correct number of factors but incorrect diagnoses loading on each factor, we generated 1,000 random variations of the 6-factor model. In each iteration, we kept the number of diagnoses per factor constant but randomly assigned diagnoses to factors. eFigure 3 displays the fit statistics and alternative indices for these 1,000 randomly generated models, as well as the fit of the preferred 6-factor model. Our preferred model demonstrated substantially better fit according to the fit statistics and had much higher factor loadings (FLs) and much lower standard deviations (SDs) than models with diagnoses randomly assigned to factors. The FLs and SDs of the random models were much closer to those from the bifactor models. Paradoxically, the mean and median standard errors (SEs) of the random models were lower than in our best-fitting model.

#### **eTable 1.** *Names and concept codes for initial set of selected diagnoses*

| Name of Condition | OMOP ID | SNOMED Code |
|---|---|---|
| Abnormal sexual function | 4237140 | 56925008 |
| Acrophobia | 4239656 | 58963008 |
| Adjustment disorder | 436677 | 17226007 |
| Adjustment disorder with anxious mood | 436075 | 47372000 |
| Adjustment disorder with depressed mood | 442306 | 57194009 |
| Adjustment disorder with disturbance of conduct | 435799 | 84984002 |
| Adjustment disorder with mixed anxiety and depressed mood | 36684319 | 782501005 |
| Adjustment disorder with mixed disturbance of emotions AND conduct | 436076 | 66381006 |
| Adjustment disorder with mixed emotional features | 433454 | 55668003 |
| Adjustment disorder with withdrawal | 437258 | 9674006 |
| Agoraphobia | 4321835 | 70691001 |
| Antisocial personality disorder | 440988 | 26665006 |
| Anxiety disorder | 442077 | 197480006 |
| Asperger's disorder | 4053178 | 23560001 |
| Attention deficit hyperactivity disorder | 438409 | 406506008 |
| Autism spectrum disorder | 439776 | 35919005 |
| Autistic disorder | 439780 | 408856003 |
| Avoidant personality disorder | 437524 | 37746008 |
| Bipolar affective disorder, current episode depression | 439254 | 191627008 |
| Bipolar affective disorder, current episode manic | 440078 | 191618007 |
| Bipolar affective disorder, current episode mixed | 435226 | 192362008 |
| Bipolar affective disorder, currently depressed, in full remission | 439251 | 191634005 |
| Bipolar affective disorder, currently depressed, mild | 439253 | 191629006 |
| Bipolar affective disorder, currently depressed, moderate | 437528 | 191630001 |
| Bipolar affective disorder, currently manic, in full remission | 439255 | 191625000 |
| Bipolar affective disorder, currently manic, mild | 441834 | 191620005 |
| Bipolar affective disorder, currently manic, moderate | 433992 | 191621009 |
| Bipolar affective disorder, currently manic, severe, with psychosis | 439256 | 191623007 |
| Bipolar affective disorder, most recent episode mixed | 35624745 | 767633005 |
| Bipolar disorder | 436665 | 13746004 |
| Bipolar disorder in full remission | 4220618 | 41836007 |
| Bipolar disorder in partial remission | 436072 | 5703000 |
| Bipolar disorder in remission | 4310821 | 85248005 |
| Bipolar disorder, most recent episode depression | 35624743 | 767631007 |
| Bipolar disorder, most recent episode manic | 35624744 | 767632000 |
| Bipolar I disorder | 432876 | 371596008 |
| Bipolar I disorder, most recent episode hypomanic | 4150985 | 31446002 |
| Bipolar I disorder, single manic episode | 432866 | 9340000 |
| Bipolar I disorder, single manic episode, in full remission | 4148842 | 3530005 |
| Bipolar I disorder, single manic episode, in remission | 4327669 | 75360000 |
| Bipolar II disorder | 4307956 | 83225003 |
| Bipolar II disorder, most recent episode major depressive | 4037669 | 16295005 |
| Bipolar type I disorder currently in full remission | 37109940 | 723903001 |
| Borderline personality disorder | 434626 | 20010003 |
| Cannabis abuse | 434327 | 37344009 |
| Cannabis dependence | 440387 | 85005007 |
| Cannabis-induced organic mental disorder | 4300092 | 77355000 |
| Chronic depressive personality disorder | 40481798 | 442057004 |
| Claustrophobia | 4058397 | 19887002 |
| Cluster A personality disorder | 4043918 | 16805009 |
| Cluster B personality disorder | 4181019 | 4306003 |
| Cluster C personality disorder | 4222618 | 83890006 |
| Conduct disorder | 443617 | 430909002 |
| Cyclothymia | 440696 | 76105009 |
| Delusional disorder | 432590 | 48500005 |
| Dependent personality disorder | 437523 | 84466009 |
| Depressed bipolar I disorder | 441836 | 49468007 |
| Depressed bipolar I disorder in full remission | 435225 | 22121000 |
| Depressed bipolar I disorder in partial remission | 4177651 | 49512000 |
| Depressed bipolar I disorder in remission | 4201739 | 53607008 |
| Disorders of initiating and maintaining sleep | 439708 | 194437008 |
| Dissociative disorder | 434889 | 44376007 |
| Disturbance in speech | 435642 | 29164008 |
| Dyspareunia | 439080 | 71315007 |
| Dysthymia | 433440 | 78667006 |
| Eating disorder | 439002 | 72366004 |
| Emotionally unstable personality disorder | 4103399 | 191765005 |

| | | |
|---|---|---|
| Explosive personality disorder | 432611 | 231527003 |
| Fear of flying | 4085062 | 247853008 |
| Fear of insects | 4023785 | 19512009 |
| Flying phobia | 4085680 | 247854002 |
| Generalized anxiety disorder | 434613 | 21897009 |
| Gilles de la Tourette's syndrome | 379782 | 5158005 |
| Gynephobia | 4012108 | 102930000 |
| Histrionic personality disorder | 440369 | 55341008 |
| Inhibited female orgasm | 442749 | 60103007 |
| Inhibited male orgasm | 444268 | 81903006 |
| Insomnia | 436962 | 193462001 |
| Intellectual disability | 40277917 | 110359009 |
| Intermittent explosive disorder | 440989 | 40987004 |
| Lack or loss of sexual desire | 443262 | 270903007 |
| Major depression, single episode | 4282096 | 36923009 |
| Major depressive disorder | 4152280 | 370143000 |
| Mania | 4333677 | 231494001 |
| Manic bipolar I disorder | 4287544 | 68569003 |
| Manic bipolar I disorder in full remission | 436086 | 30935000 |
| Manic bipolar I disorder in partial remission | 442600 | 63249007 |
| Manic bipolar I disorder in remission | 4166701 | 45479006 |
| Manic disorder, single episode | 443237 | 268619003 |
| Mild bipolar disorder | 4028027 | 13313007 |
| Mild bipolar I disorder, single manic episode | 432290 | 41552001 |
| Mild depression | 4149320 | 310495003 |
| Mild major depression | 4336957 | 87512008 |
| Mild major depression, single episode | 4195572 | 79298009 |
| Mild manic bipolar I disorder | 4215917 | 71984005 |
| Mild recurrent major depression | 4228802 | 40379007 |
| Mixed bipolar affective disorder | 439250 | 191636007 |
| Mixed bipolar affective disorder, in full remission | 439245 | 191643001 |
| Mixed bipolar affective disorder, mild | 439249 | 191638008 |
| Mixed bipolar affective disorder, moderate | 439248 | 191639000 |
| Mixed bipolar affective disorder, severe, with psychosis | 439246 | 191641004 |
| Mixed bipolar I disorder | 443906 | 16506000 |
| Mixed bipolar I disorder in full remission | 4009648 | 111485001 |
| Mixed bipolar I disorder in partial remission | 437529 | 36583000 |
| Mixed bipolar I disorder in remission | 433743 | 35481005 |
| Moderate bipolar disorder | 4194222 | 79584002 |
| Moderate bipolar I disorder, single manic episode | 440067 | 28884001 |
| Moderate depressed bipolar I disorder | 4280361 | 66631006 |
| Moderate major depression | 4307111 | 832007 |
| Moderate major depression, single episode | 4049623 | 15639000 |
| Moderate manic bipolar I disorder | 4307804 | 82998009 |
| Moderate recurrent major depression | 4077577 | 18818009 |
| Narcissistic personality disorder | 440080 | 80711002 |
| Needle phobia | 4332995 | 231501003 |
| Obsessive compulsive personality disorder | 443876 | 1376001 |
| Obsessive-compulsive disorder | 440374 | 191736004 |
| Oppositional defiant disorder | 441547 | 18941000 |
| Orgasm disorder | 4221288 | 82636008 |
| Panic disorder | 436074 | 371631005 |
| Panic disorder with agoraphobia | 4147466 | 35607004 |
| Panic disorder without agoraphobia | 4211231 | 56576003 |
| Paranoid personality disorder | 440691 | 13601005 |
| Personality disorder | 441838 | 33449004 |
| Phobic disorder | 4304010 | 386810004 |
| Posttraumatic stress disorder | 436676 | 47505003 |
| Premature ejaculation | 434319 | 44001008 |
| Psychosexual disorder | 436666 | 56627002 |
| Psychosexual dysfunction | 440068 | 268637002 |
| Psychosexual dysfunction associated with inhibited sexual excitement | 44782768 | 153151000119100 |
| Psychosis and severe depression co-occurrent and due to bipolar affective disorder | 35622934 | 765176007 |
| Psychotic disorder | 436073 | 69322001 |
| Recurrent major depression | 4282316 | 66344007 |
| Schizoaffective disorder, bipolar type | 4244078 | 38368003 |
| Schizoid personality disorder | 440085 | 52954000 |
| Schizotypal personality disorder | 434010 | 31027006 |
| Severe bipolar disorder | 4155798 | 371600003 |
| Severe bipolar disorder with psychotic features | 4195158 | 4441000 |
| Severe bipolar disorder without psychotic features | 4200385 | 53049002 |

| | | |
|---|---|---|
| Severe bipolar I disorder | 4154283 | 371599001 |
| Severe bipolar I disorder, single manic episode with psychotic features | 4220617 | 41832009 |
| Severe bipolar I disorder, single manic episode without psychotic features | 4030856 | 14495005 |
| Severe depressed bipolar I disorder | 42872413 | 261000119107 |
| Severe depressed bipolar I disorder with psychotic features | 436386 | 59617007 |
| Severe depressed bipolar I disorder without psychotic features | 442570 | 61403008 |
| Severe major depression | 42872722 | 450714000 |
| Severe major depression, single episode, without psychotic features | 441534 | 76441001 |
| Severe manic bipolar I disorder | 43020451 | 23741000119105 |
| Severe manic bipolar I disorder without psychotic features | 443797 | 162004 |
| Severe mixed bipolar I disorder | 42872412 | 271000119101 |
| Severe mixed bipolar I disorder without psychotic features | 372599 | 46229002 |
| Severe recurrent major depression without psychotic features | 435220 | 36474008 |
| Sexual desire disorder | 4186941 | 46762006 |
| Sleep disorder | 435524 | 39898005 |
| Social phobia | 440690 | 25501002 |
| Substance abuse | 4279309 | 66214007 |
| Tic disorder | 381839 | 568005 |
| Tobacco dependence syndrome | 437264 | 89765005 |
| Zoophobia | 4184002 | 54307006 |

**eTable 2**. *Frequencies and diagnostic codes for psychiatric conditions used in analyses*

| Name of Condition | N | OMOP ID | SNOMED Code | Source Vocabulary | Source Code |
|---|---|---|---|---|---|
| **Acute stress disorder** | 7,129 | 440083 | 67195008 | ICD9CM | 308.9 |
| **Adjustment disorder** | 6,532 | 436677 | 17226007 | ICD10CM | F43.20 |
| **Agoraphobia** | 181 | | | | |
| Agoraphobia | | 4321835 | 70691001 | ICD10CM | F40.00 |
| Agoraphobia without history of panic disorder | | 4189538 | 61569007 | ICD10CM | F40.02 |
| **Alcohol abuse** | 12,574 | 433753 | 15167005 | ICD10CM | F10.180 |
| **Anorexia nervosa** | 278 | 436675 | 56882008 | ICD10CM | F50.00 |
| **Antisocial personality disorder** | 461 | 440988 | 26665006 | ICD10CM | F60.2 |
| **Attention deficit hyperactivity disorder** | 4,805 | 438409 | 406506008 | ICD9CM | 314.9 |
| **Attention deficit hyperactivity disorder, combined type** | 1,012 | 4149904 | 31177006 | ICD10CM | F90.2 |
| **Attention deficit hyperactivity disorder, predominantly hyperactive impulsive type** | 393 | 4253962 | 7461003 | ICD10CM | F90.1 |
| **Attention deficit hyperactivity disorder, predominantly inattentive type** | 3,133 | 4149353 | 35253001 | ICD10CM | F90.0 |
| **Autism spectrum disorder** | 484 | | | | |
| Autism spectrum disorder | | 439776 | 35919005 | ICD9CM | 299.8 |
| Autistic disorder | | 439780 | 408856003 | ICD10CM | F84.0 |
| Asperger's disorder | | 4053178 | 23560001 | ICD10CM | F84.5 |
| **Bipolar disorder** | 10,096 | 436665 | 13746004 | ICD10CM | F31.9 |
| **Borderline personality disorder** | 1,935 | 434626 | 20010003 | ICD10CM | F60.3 |
| **Bulimia nervosa** | 375 | 438407 | 78004001 | ICD9CM | 307.51 |
| **Cannabis use** | 6,225 | | | | |
| Cannabis abuse | | 434327 | 37344009 | ICD9CM | 305.21 |
| Cannabis dependence | | 440387 | 85005007 | ICD10CM | F12.20 |
| **Claustrophobia** | 1,146 | 4058397 | 19887002 | ICD10CM | F40.240 |
| **Cocaine abuse** | 4,777 | 432303 | 78267003 | ICD10CM | F14.14 |
| **Conduct disorder** | 662 | 443617 | 430909002 | ICD10CM | F91.8 |
| **Delusional disorder** | 1,318 | 432590 | 48500005 | ICD10CM | F22 |
| **Disturbance in speech** | 2,638 | 435642 | 29164008 | ICD9CM | 784.5 |
| **Dysthymia** | 11,625 | | | | |
| Dysthymia | | 433440 | 78667006 | ICD10CM | F34.1 |
| Mild depression | | 4149320 | 310495003 | NA | NA |
| **Eating disorder** | 2,621 | 439002 | 72366004 | SNOMED | 72366004 |
| **Fear of flying** | 223 | 4085062 | 247853008 | ICD10CM | F40.243 |
| **Generalized anxiety disorder** | 19,018 | 434613 | 21897009 | ICD10CM | F41.1 |
| **Gilles de la Tourette's syndrome** | 117 | 379782 | 5158005 | ICD10CM | F95.2 |
| **Hallucinogen abuse** | 129 | 437245 | 74851005 | ICD10CM | F16.10 |
| **Intellectual disability** | 452 | 40277917 | 110359009 | ICD10CM | F79 |
| **Intermittent explosive disorder** | 241 | 440989 | 40987004 | ICD9CM | 312.34 |
| **Major depression** | 49,262 | | | | |
| Mild major depression | | 4336957 | 87512008 | SNOMED | 87512008 |
| Mild major depression, single episode | | 4195572 | 79298009 | ICD9CM | 296.21 |
| Mild recurrent major depression | | 4228802 | 40379007 | ICD10CM | F33.0 |
| Moderate major depression | | 4307111 | 832007 | SNOMED | 832007 |
| Recurrent major depression | | 4282316 | 66344007 | ICD9CM | 296.3 |
| Severe major depression | | 42872722 | 450714000 | SNOMED | 450714000 |
| Severe major depression, single episode, without psychotic features | | 441534 | 76441001 | ICD9CM | 296.23 |
| Severe recurrent major depression without psychotic features | | 435220 | 36474008 | ICD10CM | F33.2 |
| **Narcissistic personality disorder** | 178 | 440080 | 80711002 | ICD9CM | 301.81 |
| **Obsessive compulsive personality disorder** | 183 | 443876 | 1376001 | ICD9CM | 301.4 |
| **Obsessive-compulsive disorder** | 1,638 | 440374 | 191736004 | ICD10CM | F42 |
| **Opioid abuse** | 4,383 | 438130 | 5602001 | ICD10CM | F11.10 |
| **Oppositional defiant disorder** | 255 | 441547 | 18941000 | ICD9CM | 313.81 |
| **Panic disorder** | 8,608 | | | | |
| Panic disorder | | 436074 | 371631005 | SNOMED | 371631005 |
| Panic disorder with agoraphobia | | 4147466 | 35607004 | ICD9CM | 300.21 |
| Panic disorder without agoraphobia | | 4211231 | 56576003 | ICD10CM | F41.0 |
| **Posttraumatic stress disorder** | 10,837 | 436676 | 47505003 | ICD10CM | F43.10 |
| **Schizoaffective disorder** | 2,473 | 4286201 | 68890003 | ICD10CM | F25.9 |
| **Schizophrenia** | 3,154 | 435783 | 58214004 | ICD10CM | F20.89 |
| **Sedative abuse** | 347 | 442601 | 64386003 | ICD10CM | F13.11 |
| **Sleep disorder (used as an external correlate)** | 12,985 | 435524 | 39898005 | ICD10CM | G47.9 |
| **Social phobia** | 947 | 440690 | 25501002 | ICD10CM | F40.10 |
| **Tic disorder** | 228 | 381839 | 568005 | ICD10CM | F95.9 |
| **Tobacco dependence syndrome** | 21,187 | 437264 | 89765005 | ICD9CM | 305.1 |

**eTable 3.** *All of Us sample characteristics across race-ethnicity*

| | White (N=72,806) | African American (N=24,952) | Asian (N=2,146) | Multiracial (N=2,235) | Hispanic only (N=18,978) | Hispanic (N=21,693) | Non-Hispanic (N=106,270) | Other (N=783) | Not reported (N=6,063) | Total (N=127,963) |
|---|---|---|---|---|---|---|---|---|---|---|
| **Sex Assigned at Birth** | | | | | | | | | | |
| Female | 44,587 (61.2%) | 15,035 (60.3%) | 1,328 (61.9%) | 1,509 (67.5%) | 12,662 (66.7%) | 14,452 (66.6%) | 63,279 (59.5%) | 400 (51.1%) | 2,210 (36.5%) | 77,731 (60.7%) |
| Male | 27,514 (37.8%) | 9,479 (38.0%) | 799 (37.2%) | 691 (30.9%) | 6,129 (32.3%) | 7,029 (32.4%) | 39,512 (37.2%) | 367 (46.9%) | 1,562 (25.8%) | 46,541 (36.4%) |
| No matching concept | 42 (0.1%) | 43 (0.2%) | 4 (0.2%) | 3 (0.1%) | 15 (0.1%) | 17 (0.1%) | 2,160 (2.0%) | 1 (0.1%) | 2,069 (34.1%) | 2,177 (1.7%) |
| Not male, not female, prefer not to answer, or skipped | 663 (0.9%) | 395 (1.6%) | 15 (0.7%) | 32 (1.4%) | 172 (0.9%) | 195 (0.9%) | 1,319 (1.2%) | 15 (1.9%) | 222 (3.7%) | 1,514 (1.2%) |
| **Gender** | | | | | | | | | | |
| Female | 44,222 (60.7%) | 14,988 (60.1%) | 1,311 (61.1%) | 1,458 (65.2%) | 12,572 (66.2%) | 14,328 (66.0%) | 62,760 (59.1%) | 395 (50.4%) | 2,142 (35.3%) | 77,088 (60.2%) |
| Male | 27,289 (37.5%) | 9,443 (37.8%) | 794 (37.0%) | 689 (30.8%) | 6,067 (32.0%) | 6,952 (32.0%) | 39,217 (36.9%) | 360 (46.0%) | 1,527 (25.2%) | 46,169 (36.1%) |
| Not man only, not woman only, prefer not to answer, or skipped | 1,295 (1.8%) | 521 (2.1%) | 41 (1.9%) | 88 (3.9%) | 339 (1.8%) | 413 (1.9%) | 4293 (4.0%) | 28 (3.6%) | 2,394 (39.5%) | 4,706 (3.7%) |
| **Sexual Orientation** | | | | | | | | | | |
| Heterosexual | 64,718 (88.9%) | 21,512 (86.2%) | 1,881 (87.7%) | 1,790 (80.1%) | 16,741 (88.2%) | 18,929 (87.3%) | 91,515 (86.1%) | 684 (87.4%) | 3,118 (51.4%) | 110,444 (86.3%) |
| All other orientations | 8,088 (11.1%) | 3,440 (13.8%) | 265 (12.3%) | 445 (19.9%) | 2,237 (11.8%) | 2,764 (12.7%) | 14,755 (13.9%) | 99 (12.6%) | 2,945 (48.6%) | 17,519 (13.7%) |
| **Household Income ($/year)** | | | | | | | | | | |
| Less than 10000 | 6,916 (9.5%) | 8,224 (33.0%) | 151 (7.0%) | 374 (16.7%) | 4,132 (21.8%) | 4,686 (21.6%) | 15,971 (15.0%) | 100 (12.8%) | 760 (12.5%) | 20,657 (16.1%) |
| 10000 to 25000 | 8,639 (11.9%) | 4,591 (18.4%) | 171 (8.0%) | 339 (15.2%) | 3,029 (16.0%) | 3,427 (15.8%) | 14,017 (13.2%) | 107 (13.7%) | 568 (9.4%) | 17,444 (13.6%) |
| 25000 to 35000 | 5,518 (7.6%) | 1,823 (7.3%) | 104 (4.8%) | 172 (7.7%) | 1,398 (7.4%) | 1,634 (7.5%) | 7,700 (7.2%) | 44 (5.6%) | 275 (4.5%) | 9,334 (7.3%) |
| 35000 to 50000 | 6,614 (9.1%) | 1,462 (5.9%) | 173 (8.1%) | 199 (8.9%) | 1,166 (6.1%) | 1,389 (6.4%) | 8,573 (8.1%) | 66 (8.4%) | 282 (4.7%) | 9,962 (7.8%) |
| 50000 to 75000 | 9,651 (13.3%) | 1,307 (5.2%) | 221 (10.3%) | 209 (9.4%) | 998 (5.3%) | 1,267 (5.8%) | 11,485 (10.8%) | 78 (10.0%) | 288 (4.8%) | 12,752 (10.0%) |
| 75000 to 100000 | 7,567 (10.4%) | 614 (2.5%) | 219 (10.2%) | 168 (7.5%) | 510 (2.7%) | 687 (3.2%) | 8,683 (8.2%) | 52 (6.6%) | 240 (4.0%) | 9,370 (7.3%) |
| 100000 to 150000 | 9,084 (12.5%) | 467 (1.9%) | 297 (13.8%) | 171 (7.7%) | 446 (2.4%) | 626 (2.9%) | 10,113 (9.5%) | 57 (7.3%) | 217 (3.6%) | 10,739 (8.4%) |
| 150000 to 200000 | 3,967 (5.4%) | 148 (0.6%) | 145 (6.8%) | 75 (3.4%) | 133 (0.7%) | 213 (1.0%) | 4,370 (4.1%) | 35 (4.5%) | 80 (1.3%) | 4,583 (3.6%) |
| More than 200000 | 5,254 (7.2%) | 139 (0.6%) | 230 (10.7%) | 119 (5.3%) | 166 (0.9%) | 245 (1.1%) | 5,845 (5.5%) | 48 (6.1%) | 134 (2.2%) | 6,090 (4.8%) |
| Not reported | 9,596 (13.2%) | 6,177 (24.8%) | 435 (20.3%) | 409 (18.3%) | 7,000 (36.9%) | 7,519 (34.7%) | 19,513 (18.4%) | 196 (25.0%) | 3,219 (53.1%) | 27,032 (21.1%) |
| **Education** | | | | | | | | | | |
| Less than a high school degree or equivalent | 2,791 (3.8%) | 3,930 (15.8%) | 40 (1.9%) | 140 (6.3%) | 5,629 (29.7%) | 5,894 (27.2%) | 7,074 (6.7%) | 34 (4.3%) | 404 (6.7%) | 12,968 (10.1%) |
| Highest Grade: Twelve Or GED | 11,736 (16.1%) | 8,275 (33.2%) | 150 (7.0%) | 398 (17.8%) | 5,059 (26.7%) | 5,635 (26.0%) | 21,030 (19.8%) | 137 (17.5%) | 910 (15.0%) | 26,665 (20.8%) |
| Highest Grade: College One to Three | 20,979 (28.8%) | 7,555 (30.3%) | 320 (14.9%) | 742 (33.2%) | 4,632 (24.4%) | 5,595 (25.8%) | 29,867 (28.1%) | 168 (21.5%) | 1,066 (17.6%) | 35,462 (27.7%) |
| College graduate or advanced degree | 36,566 (50.2%) | 4,078 (16.3%) | 1,602 (74.7%) | 903 (40.4%) | 3,087 (16.3%) | 3,965 (18.3%) | 43,947 (41.4%) | 419 (53.5%) | 1,257 (20.7%) | 47,912 (37.4%) |
| Not reported | 734 (1.0%) | 1,114 (4.5%) | 34 (1.6%) | 52 (2.3%) | 571 (3.0%) | 604 (2.8%) | 4,352 (4.1%) | 25 (3.2%) | 2,426 (40.0%) | 4,956 (3.9%) |
| **Marital Status** | | | | | | | | | | |
| Married | 35,585 (48.9%) | 4,446 (17.8%) | 1,054 (49.1%) | 661 (29.6%) | 6,201 (32.7%) | 6,958 (32.1%) | 42,587 (40.1%) | 336 (42.9%) | 1,262 (20.8%) | 49,545 (38.7%) |
| Living With Partner | 4,198 (5.8%) | 1,460 (5.9%) | 111 (5.2%) | 207 (9.3%) | 1,618 (8.5%) | 1,910 (8.8%) | 5,938 (5.6%) | 34 (4.3%) | 220 (3.6%) | 7,848 (6.1%) |
| Divorced | 12,266 (16.8%) | 4,432 (17.8%) | 187 (8.7%) | 305 (13.6%) | 3,306 (17.4%) | 3,709 (17.1%) | 17,582 (16.5%) | 113 (14.4%) | 682 (11.2%) | 2,1291 (16.6%) |

| | | | | | | | | | | |
|---|---|---|---|---|---|---|---|---|---|---|
| Never Married | 13,581 (18.7%) | 10,111 (40.5%) | 642 (29.9%) | 839 (37.5%) | 4,781 (25.2%) | 5,766 (26.6%) | 25,412 (23.9%) | 228 (29.1%) | 996 (16.4%) | 31,178 (24.4%) |
| Separated | 1,568 (2.2%) | 1,657 (6.6%) | 32 (1.5%) | 68 (3.0%) | 1,432 (7.5%) | 1,537 (7.1%) | 3,402 (3.2%) | 19 (2.4%) | 163 (2.7%) | 4,939 (3.9%) |
| Widowed | 4,647 (6.4%) | 1,631 (6.5%) | 76 (3.5%) | 85 (3.8%) | 983 (5.2%) | 1,066 (4.9%) | 6,659 (6.3%) | 31 (4.0%) | 272 (4.5%) | 7,725 (6.0%) |
| Not reported | 961 (1.3%) | 1,215 (4.9%) | 44 (2.1%) | 70 (3.1%) | 657 (3.5%) | 747 (3.4%) | 4,690 (4.4%) | 22 (2.8%) | 2,468 (40.7%) | 5,437 (4.2%) |

#### **eTable 4**. *Fit and alternative indices for confirmatory and exploratory models*

| | | | | | | | Alternative Indices | | | Errors |
|---|---|---|---|---|---|---|---|---|---|---|
| **Model** | $\chi^2$ | **df** | **CFI** | **TLI** | **RMSEA** | **SRMR** | **FL** | **SD** | **SE** | |
| **1.** General factor | 43,769 | 702 | .807 | .797 | 0.022 | 0.13 | .493 | .167 | .014 | |
| ***Correlated factors (I=Internalizing, E=Externalizing, T=Thought problems, S=Substance use, N=Neurodevelopmental disorders, F=Fear, D = Distress)*** | | | | | | | | | | |
| **2.** I, E, T | 24,669 | 591 | .890 | .883 | .018 | .123 | .556 | .182 | .015 | |
| **3.** I, E, T, S | 18,888 | 588 | .916 | .910 | .016 | .115 | .631 | .194 | .015 | |
| **4.** I, E, T, N | 25,128 | 696 | .891 | .884 | .017 | .120 | .567 | .170 | .016 | |
| **5.** I, E, T, S, N | 19,187 | 692 | .917 | .911 | .014 | .108 | .604 | .177 | .016 | Yes |
| **6.** F, D, E, T | 24,535 | 588 | .891 | .883 | .018 | .123 | .572 | .182 | .015 | |
| **7.** F, D, E, T, S | 18,750 | 584 | .917 | .910 | .016 | .114 | .644 | .192 | .016 | |
| **8.** F, D, E, T, N | 24,895 | 690 | .892 | .884 | .017 | .119 | .587 | .278 | .020 | |
| **9.** F, D, E, T, S, N | 19,050 | 687 | .918 | .911 | .014 | .107 | .613 | .175 | .016 | Yes |
| ***Tests of the placement of ambiguously classified disorders*** | | | | | | | | | | |
| ***ADHD and its three subtypes*** | | | | | | | | | | |
| *Placement in model 9: ADHD-HI & ADHD-C on E; ADHD-IN on N* | | | | | | | | | | |
| **9.1.** Six correlated factors (ADHD on E only) | 17,449 | 614 | .923 | .917 | .015 | .101 | .655 | .181 | .016 | |
| **9.2.** Six correlated factors (ADHD on N only) | 17,329 | 614 | .924 | .917 | .015 | .101 | .615 | .182 | .016 | |
| **9.3.** Six correlated factors (ADHD on E and N) | 17,293 | 613 | .924 | .917 | .015 | .101 | .622 | .192 | .017 | |
| **9.4.** Six correlated factors (ADHD-HI and ADHD-C on E; ADHD-IN and ADHD-C on N) | 18,699 | 686 | .919 | .913 | .014 | .105 | .619 | .215 | .019 | |
| **9.5.** Six correlated factors (ADHD-HI on E; ADHD-IN and ADHD-C on N. Correlated residuals between ADHD-HI and ADHD-IN as well as ADHD-HI and ADHD-C) | 18,299 | 685 | .921 | .915 | .014 | .102 | .616 | .176 | .017 | |
| **9.6.** Six correlated factors (ADHD-C on E; ADHD-HI and ADHD-IN on N. Correlated residuals between ADHD-C and ADHD-HI as well as ADHD-C and ADHD-IN) | 18,322 | 685 | .921 | .915 | .014 | .103 | .622 | .177 | .017 | |
| ***Eating Disorder (ED), Anorexia Nervosa (AN), and Bulimia Nervosa (BN)*** | | | | | | | | | | |
| *Placement in model 9.5: ED on T* | | | | | | | | | | |
| **9.5.1.** Six correlated factors (ED on D only) | 17,550 | 685 | .925 | .918 | .014 | .101 | .614 | .174 | .017 | |
| **9.5.2.** Six correlated factors (ED on D and T) | 17,530 | 684 | .925 | .918 | .014 | .101 | .614 | .201 | .017 | |
| **9.5.3.** Six correlated factors (AN on D and T; BN on D and E) | 17,880 | 721 | .924 | .917 | .014 | .103 | .613 | .211 | .019 | |
| **9.5.4.** Six correlated factors (AN on D; BN on D and E) | 17,880 | 723 | .924 | .918 | .014 | .103 | .614 | .168 | .017 | |
| **9.5.5.** Six correlated factors (AN on T; BN on E) | 18,299 | 723 | .922 | .916 | .014 | .107 | .615 | .166 | .017 | |
| ***Obsessive-Compulsive Disorder (OCD) & Obsessive-Compulsive Personality Disorder (OCPD)*** | | | | | | | | | | |
| *Placement in model 9.5: OCD & OCPD on T* | | | | | | | | | | |
| **9.5.6.** Six correlated factors (OCD and OCPD on F) | 16,740 | 685 | .928 | .922 | .014 | .097 | .654 | .178 | .017 | |
| **9.5.7.** Six correlated factors (OCD and OCPD on N) | 16,765 | 685 | .928 | .922 | .014 | .098 | .611 | .182 | .016 | |
| **9.5.8.** Six correlated factors (OCD and OCPD on F and N) | 16,418 | 683 | .930 | .924 | .013 | .096 | .606 | .183 | .018 | |
| **9.5.9.** Six correlated factors (OCD and OCPD on D) | 16,739 | 685 | .928 | .922 | .014 | .097 | .637 | .177 | .017 | |
| ***Borderline Personality Disorder (BPD)*** | | | | | | | | | | |
| *Placement in model 9.5: BPD on T* | | | | | | | | | | |
| **9.5.10.** Six correlated factors (BPD on E) | 17,244 | 685 | .926 | .920 | .014 | .101 | .611 | .176 | .016 | |
| **9.5.11.** Six correlated factors (BPD on D) | 17,775 | 685 | .924 | .917 | .014 | .103 | .619 | .179 | .017 | |
| **9.5.12.** Six correlated factors (BPD on D and E) | 17,051 | 684 | .927 | .921 | .014 | .101 | .617 | .172 | .017 | |
| **9.5.13.** Six correlated factors (BPD on D and T) | 16,773 | 684 | .928 | .922 | .014 | .102 | .615 | .177 | .017 | |
| **9.5.14.** Six correlated factors (BPD on E and T) | 17,238 | 684 | .926 | .920 | .014 | .101 | .610 | .188 | .017 | |
| ***Tic and Tourette's (TS)*** | | | | | | | | | | |
| *Placement in model 9.5: Tic and TS on N* | | | | | | | | | | |
| **9.5.15.** Six correlated factors (TIC and TS on D) | 18,548 | 685 | .920 | .914 | .014 | .105 | .619 | .185 | .016 | |
| **9.5.16.** Six correlated factors (TIC and TS on N and D) | 18,275 | 683 | .921 | .915 | .014 | .103 | .634 | .237 | .019 | |
| ***Antisocial personality disorder (ASPD)*** | | | | | | | | | | |
| *Placement in model 9.5: ASPD on S* | | | | | | | | | | |
| **9.5.17.** Six correlated factors (ASPD on E and S) | 17,904 | 684 | .923 | .917 | .014 | .101 | .614 | .172 | .017 | |
| ***Additional Correlated Residuals (~~)*** | | | | | | | | | | |
| **9.5.18.** Six correlated factors (SCZ ~~ SCZAFF) | 16,761 | 684 | .928 | .922 | .014 | .100 | .614 | .172 | .017 | |
| **9.5.19.** Six correlated factors (OCD ~~ OCPD) | 17,929 | 684 | .923 | .916 | .014 | .100 | .617 | .178 | .017 | |
| **9.5.20.** Six correlated factors (MDD ~~ dysthymia) | 17,863 | 684 | .923 | .917 | .014 | .102 | .615 | .177 | .017 | |
| **9.5.21.** Six correlated factors (TIC ~~ TS) | 18,131 | 684 | .922 | .916 | .014 | .100 | .615 | .180 | .017 | |
| **9.5.22.** Six correlated factors (CD ~~ ODD) | 18,172 | 684 | .922 | .915 | .014 | .100 | .607 | .172 | .017 | Yes |
| **9.5.23.** Six correlated factors (OCD ~~ TS) | 18,243 | 684 | .922 | .915 | .014 | .102 | .616 | .176 | .017 | |
| **9.5.24.** Six correlated factors (OCD ~~ TIC) | 18,254 | 684 | .921 | .915 | .014 | .102 | .616 | .176 | .017 | |

| | | | | | | | | | |
|---|---|---|---|---|---|---|---|---|---|
| **9.5.25.** Six correlated factors (PANIC ~~ AGOR) | 18,270 | 684 | .921 | .915 | .014 | .102 | .616 | .177 | .017 |
| **9.5.26.** Six correlated factors (SCZ ~~ SCZAFF; OCD ~~ OCPD; MDD ~~ dysthymia) | 15,962 | 682 | .932 | .926 | .013 | .098 | .617 | .174 | .017 |
| ***Models Based on Modification Indices*** | | | | | | | | | |
| ***PTSD** - Previous placement in Model 9.5.26: PTSD on D* | | | | | | | | | |
| **9.5.26.1.** Six correlated factors (PTSD on T) | 14,537 | 682 | .938 | .933 | .013 | .097 | .616 | .169 | .017 |
| **9.5.26.2.** Six correlated factors (PTSD on T and D) | 13,428 | 681 | .943 | .938 | .012 | .097 | .614 | .175 | .017 |
| ***OCD** - Previous placement in Model 9.5.26: OCD on D* | | | | | | | | | |
| **9.5.26.2.1.** Six correlated factors (OCD on T and D) | 12,147 | 680 | .949 | .944 | .011 | .094 | .611 | .188 | .017 |
| ***BPD** - Previous placement in Model 9.5.26: BPD on T* | | | | | | | | | |
| **9.5.26.2.1.1.** Six correlated factors (BPD on T and D) | 10,858 | 679 | .954 | .950 | .011 | .093 | .605 | .185 | .017 |
| ***ED** - Previous placement in Model 9.5.26: ED on T* | | | | | | | | | |
| **9.5.26.2.1.1.1.** Six correlated factors (ED on T and D) | 9,469 | 678 | .961 | .957 | .010 | .091 | .598 | .202 | .017 |

*Note*. CFI – comparative fit index, TLI – tucker-lewis index, RMSEA - root mean square error of approximation, SRMR – standardized root mean squared residual, FL – median factor loading, SD – standard deviation of loadings, SE – mean standard error. ADHD-HI – ADHD hyperactive-impulsive subtype, ADHD-IN – ADHD inattentive subtype, ADHD-C = ADHD combined subtype, SCZ – schizophrenia, SCZAFF – schizoaffective, CD – conduct disorder, ODD – oppositional defiant disorder, PANIC – panic disorder, AGOR – agoraphobia. Model in bold is our preferred best-fitting model.

**eTable 5**. *Alternative indices for fear and distress diagnoses across confirmatory models*

| | Alternative Indices Fear Diagnoses | | | Alternative Indices Distress Diagnoses | | |
|---|---|---|---|---|---|---|
| **Model** | **FL** | **SD** | **SE** | **FL** | **SD** | **SE** |
| **2.** I, E, T | .525 | .270 | .020 | .464 | .154 | .013 |
| **3.** I, E, T, S | .530 | .267 | .020 | .468 | .150 | .013 |
| **4.** I, E, T, N | .533 | .270 | .020 | .549 | .124 | .015 |
| **5.** I, E, T, S, N | .535 | .268 | .020 | .545 | .121 | .015 |
| **6.** F, D, E, T | .580 | .301 | .022 | .461 | .153 | .013 |
| **7.** F, D, E, T, S | .588 | .298 | .022 | .465 | .149 | .022 |
| **8.** F, D, E, T, N | .588 | .300 | .022 | .463 | .434 | .020 |
| **9.** F, D, E, T, S, N | .591 | .298 | .022 | .547 | .140 | .007 |

*Note*. F = fear, D = distress, E = externalizing, S = substance use, T = thought problems, N = neurodevelopmental disorders. FL = median factor loading, SD = standard deviation of loadings, SE = mean standard error.

**eTable 6.** *Fit and alternative indices for bifactor and modified bifactor versions of best fitting models*

| | | | | | | | Alternative Indices | | | Errors |
|---|---|---|---|---|---|---|---|---|---|---|
| **Model** | **χ²** | **df** | **CFI** | **TLI** | **RMSEA** | **SRMR** | **FL** | **SD** | **SE** | |
| **Bifactor (Orthogonal specific factors)** | | | | | | | | | | |
| **2.** I, E, T | 12,161 | 558 | .947 | .940 | .013 | .097 | .298 | .369 | .018 | |
| **3.** I, E, T, S | 12,169 | 558 | .947 | .940 | .013 | .094 | .429 | .314 | .019 | |
| **4.** I, E, T, N | 14,789 | 663 | .937 | .929 | .013 | .098 | .363 | .343 | .020 | |
| **5.** I, E, T, S, N | 15,459 | 663 | .934 | .926 | .013 | .099 | .457 | .313 | .021 | |
| **6.** F, D, E, T | 17,249 | 558 | .924 | .914 | .015 | .101 | .305 | .359 | .018 | |
| **7.** F, D, E, T, S | 16,993 | 558 | .925 | .915 | .015 | .099 | .389 | .317 | .019 | |
| **8.** F, D, E, T, N | 19,486 | 661 | .916 | .906 | .015 | .100 | .359 | .322 | .022 | |
| **9.** F, D, E, T, S, N | 20,171 | 663 | .913 | .903 | .015 | .102 | .421 | .301 | .031 | |
| **9.5.** Six correlated factors (ADHD-HI on E; ADHD-IN and ADHD-C on N) | 17,354 | 661 | .925 | .916 | .014 | .097 | .398 | .295 | .021 | |
| **9.5.26.** Six correlated factors (9.5 + SCZ ~~ SCZAFF; OCD ~~ OCPD; MDD ~~ dysthymia) | 17,069 | 658 | .927 | .917 | .014 | .095 | .416 | .271 | .029 | |
| **9.5.26.2.1.1.1.** Six correlated factors (9.5.27 + PTSD on T and D; OCD on T and D; BPD on T and D; ED on D and T) | 16,360 | 654 | .930 | .92 | .014 | .095 | .379 | .280 | .024 | Yes |
| **Modified Bifactor (Correlated specific factors)** | | | | | | | | | | |
| **2.** I, E, T | 7,005 | 555 | .971 | .967 | .010 | .081 | .397 | .314 | .018 | |
| **3.** I, E, T, S | 5,756 | 552 | .976 | .973 | .009 | .076 | .420 | .279 | .019 | |
| **4.** I, E, T, N | 7,466 | 657 | .970 | .966 | .009 | .080 | .425 | .298 | .019 | |
| **5.** I, E, T, S, N | 6,351 | 653 | .975 | .971 | .008 | .077 | .419 | .276 | .020 | Yes |
| **6.** F, D, E, T | 6,922 | 552 | .971 | .967 | .009 | .080 | .401 | .315 | .018 | |
| **7.** F, D, E, T, S | 5,681 | 548 | .977 | .973 | .009 | .075 | .412 | .280 | .019 | |
| **8.** F, D, E, T, N | 7,375 | 651 | .970 | .966 | .009 | .080 | .415 | .304 | .019 | |
| **9.** F, D, E, T, S, N | 6,286 | 648 | .975 | .971 | .008 | .077 | .420 | .276 | .020 | Yes |
| **9.5.** Six correlated factors (ADHD-HI on E; ADHD-IN and ADHD-C on N) | 5,767 | 646 | .977 | .974 | .008 | .072 | .402 | .257 | .021 | |
| **9.5.26.** Six correlated factors (9.5 + SCZ ~~ SCZAFF; OCD ~~ OCPD; MDD ~~ dysthymia) | 5,095 | 643 | .980 | .977 | .007 | .070 | .400 | .250 | .022 | |
| **9.5.26.2.1.1.1.** Six correlated factors (9.5.27 + PTSD on T and D; OCD on T and D; BPD on T and D; ED on D and T) | 4,912 | 639 | .981 | .978 | .007 | .069 | .395 | .396 | .051 | |

*Note*. CFI – comparative fit index, TLI – tucker-lewis index, RMSEA - root mean square error of approximation, SRMR – standardized root mean squared residual, FL – median factor loading, SD – standard deviation of loadings, SE – mean standard error. ADHD-HI – ADHD hyperactive-impulsive subtype, ADHD-IN – ADHD inattentive subtype, ADHD-C = ADHD combined subtype, SCZ – schizophrenia, SCZAFF – schizoaffective, CD – conduct disorder, MDD – major depressive disorder, OCD – obsessive-compulsive disorder, BPD – borderline personality disorder, ED – eating disorder.

**eTable 7**. *Comparisons among factors in Model 9.5 in their alternative indices*

| Index / Factor | Mean FLs | Median FLs | SD of FLs | Mean SEs of FLs | Median SEs of FLs |
|---|---|---|---|---|---|
| **Externalizing** | .688 | .700 | .110 | .027 | .028 |
| **Substance Use** | .692 | .693 | .096 | .012 | .007 |
| **Thought Problems** | .641 | .670 | .169 | .012 | .011 |
| **Fear** | .431 | .591 | .297 | .022 | .018 |
| **Distress** | .520 | .547 | .014 | .007 | .006 |
| **NDD** | .587 | .616 | .135 | .026 | .023 |

**eTable 8**. *Split-half replication of all models*

| Model | $\chi^2$ | df | CFI | TLI | RMSEA | SRMR | Alternative Indices | | |
|---|---|---|---|---|---|---|---|---|---|
| | | | | | | | **FL** | **SD** | **SE** |
| **1.** General factor | 43,769 | 702 | 0.807 | 0.797 | 0.022 | 0.127 | .493 | .17 | .01 |
| **a. First random half** | 22,695 | 702 | 0.806 | 0.795 | 0.022 | 0.13 | .468 | .160 | .02 |
| **b. Second random half** | 22,115 | 702 | 0.808 | 0.798 | 0.022 | 0.13 | .505 | .165 | .02 |
| **2.** I, E, T | 24,669 | 591 | 0.89 | 0.883 | 0.018 | 0.123 | 0.56 | 0.18 | 0.01 |
| **a. First random half** | 12,381 | 591 | 0.893 | 0.886 | 0.018 | 0.124 | 0.54 | 0.17 | 0.02 |
| **b. Second random half** | 13,169 | 591 | 0.885 | 0.877 | 0.018 | 0.13 | 0.57 | 0.18 | 0.02 |
| **3.** I, E, T, S | 18,888 | 588 | 0.916 | 0.91 | 0.016 | 0.115 | 0.63 | 0.19 | 0.02 |
| **a. First random half** | 9,636 | 588 | 0.918 | 0.912 | 0.016 | 0.116 | 0.64 | 0.18 | 0.02 |
| **b. Second random half** | 10,159 | 588 | 0.912 | 0.906 | 0.016 | 0.121 | 0.64 | 0.19 | 0.02 |
| **4.** I, E, T, N | 25,128 | 696 | 0.891 | 0.884 | 0.017 | 0.12 | 0.57 | 0.17 | 0.02 |
| **a. First random half** | 12,774 | 696 | 0.893 | 0.886 | 0.016 | 0.123 | 0.55 | 0.16 | 0.02 |
| **b. Second random half** | 13,398 | 696 | 0.886 | 0.879 | 0.017 | 0.125 | 0.58 | 0.17 | 0.02 |
| **5.** I, E, T, S, N | 19,187 | 692 | 0.917 | 0.911 | 0.014 | 0.108 | 0.6 | 0.18 | 0.02 |
| **a. First random half** | 10,035 | 692 | 0.917 | 0.912 | 0.015 | 0.11 | 0.59 | 0.17 | 0.02 |
| **b. Second random half** | 10,327 | 692 | 0.914 | 0.908 | 0.015 | 0.113 | 0.62 | 0.18 | 0.02 |
| **6.** F, D, E, T | 24,535 | 588 | 0.891 | 0.883 | 0.018 | 0.123 | 0.57 | 0.18 | 0.02 |
| **a. First random half** | 12,320 | 588 | 0.894 | 0.886 | 0.018 | 0.124 | 0.54 | 0.17 | 0.02 |
| **b. Second random half** | 13,092 | 588 | 0.886 | 0.877 | 0.018 | 0.129 | 0.57 | 0.18 | 0.02 |
| **7.** F, D, E, T, S | 18,750 | 584 | 0.917 | 0.91 | 0.016 | 0.114 | 0.64 | 0.19 | 0.02 |
| **a. First random half** | 9,571 | 584 | 0.919 | 0.912 | 0.016 | 0.115 | 0.65 | 0.18 | 0.02 |
| **b. Second random half** | 9,571 | 584 | 0.919 | 0.912 | 0.016 | 0.115 | 0.65 | 0.18 | 0.02 |
| **8.** F, D, E, T, N | 24,895 | 690 | 0.892 | 0.884 | 0.017 | 0.119 | 0.59 | 0.28 | 0.02 |
| **a. First random half** | 12,695 | 690 | 0.894 | 0.886 | 0.016 | 0.122 | 0.55 | 0.24 | 0.03 |
| **b. Second random half** | 13,272 | 690 | 0.887 | 0.879 | 0.017 | 0.124 | 0.58 | 0.28 | 0.03 |
| **9.** F, D, E, T, S, N | 19,050 | 687 | 0.918 | 0.911 | 0.014 | 0.107 | 0.61 | 0.17 | 0.02 |
| **a. First random half** | 9,972 | 687 | 0.918 | 0.911 | 0.015 | 0.109 | 0.62 | 0.17 | 0.02 |
| **b. Second random half** | 10,240 | 687 | 0.914 | 0.908 | 0.015 | 0.112 | 0.62 | 0.17 | 0.02 |
| **9.1.** Six correlated factors (ADHD on E only) | 17,449 | 614 | 0.923 | 0.917 | 0.015 | 0.101 | 0.65 | 0.18 | 0.02 |
| **a. First random half** | 9,113 | 614 | 0.923 | 0.917 | 0.015 | 0.104 | 0.61 | 0.17 | 0.02 |
| **b. Second random half** | 9,275 | 614 | 0.921 | 0.914 | 0.015 | 0.106 | 0.65 | 0.18 | 0.02 |
| **9.2.** Six correlated factors (ADHD on N only) | 17,329 | 614 | 0.924 | 0.917 | 0.015 | 0.101 | 0.61 | 0.18 | 0.02 |
| **a. First random half** | 9,053 | 614 | 0.924 | 0.918 | 0.015 | 0.104 | 0.6 | 0.17 | 0.02 |
| **b. Second random half** | 9,187 | 614 | 0.921 | 0.915 | 0.015 | 0.105 | 0.61 | 0.18 | 0.02 |
| **9.3.** Six correlated factors (ADHD on E and N) | 17,293 | 613 | 0.924 | 0.917 | 0.015 | 0.101 | 0.62 | 0.19 | 0.02 |
| **a. First random half** | 9,036 | 613 | 0.924 | 0.918 | 0.015 | 0.104 | 0.6 | 0.19 | 0.02 |
| **b. Second random half** | 9,175 | 613 | 0.922 | 0.915 | 0.015 | 0.105 | 0.62 | 0.19 | 0.02 |
| **9.4.** Six correlated factors (Hyp-Imp and Combined on E; Inattentive and Combined on N) | 18,699 | 686 | 0.919 | 0.913 | 0.014 | 0.105 | 0.62 | 0.22 | 0.02 |
| **a. First random half** | 9,821 | 686 | 0.919 | 0.913 | 0.014 | 0.109 | 0.64 | 0.21 | 0.03 |
| **b. Second random half** | 10,009 | 686 | 0.917 | 0.91 | 0.015 | 0.109 | 0.62 | 0.22 | 0.03 |
| **9.5.** Six correlated factors (Hyp-Imp on E; Inattentive and COMB on N. Correlated residuals between Hyp-Imp and Inattentive as well as Hyp-Imp and Combined) | 18,299 | 685 | .921 | .915 | .014 | .1 | .616 | .176 | .02 |
| **a. First random half** | 9,579 | 685 | .921 | .915 | .014 | .107 | .640 | .170 | .02 |
| **b. Second random half** | 9,783 | 685 | .919 | .912 | .014 | .107 | .640 | .180 | .02 |
| **9.6.** Six correlated factors (Combined on E; Hyp-Imp and Inattentive on N. Correlated residuals between Combined and Hyp-Imp as well as Combined and Inattentive) | 18,322 | 685 | 0.921 | 0.915 | 0.014 | 0.103 | 0.62 | 0.18 | 0.02 |
| **a. First random half** | 9,562 | 685 | 0.921 | 0.915 | 0.014 | 0.105 | 0.61 | 0.17 | 0.02 |
| **b. Second random half** | 9,868 | 685 | 0.918 | 0.911 | 0.014 | 0.108 | 0.64 | 0.18 | 0.02 |
| **9.5.1.** Six correlated factors (ED on D only) | 17,550 | 685 | .925 | .918 | .014 | .1 | .614 | .174 | .02 |
| **a. First random half** | 9,176 | 685 | .925 | .919 | .014 | .106 | .640 | .170 | .02 |
| **b. Second random half** | 9,437 | 685 | .922 | .915 | .014 | .106 | .640 | .170 | .02 |
| **9.5.2.** Six correlated factors (ED on D and T) | 17,530 | 684 | .925 | .918 | .014 | .1 | .614 | .201 | .02 |
| **a. First random half** | 9,165 | 684 | .925 | .919 | .014 | .106 | .620 | .190 | .02 |
| **b. Second random half** | 9,428 | 684 | .922 | .915 | .014 | .106 | .630 | .200 | .02 |
| **9.5.3.** Six correlated factors (AN on D and T; BN on D and E) | 17,880 | 721 | .924 | .917 | .014 | .1 | .613 | .211 | .02 |
| **a. First random half** | 9,291 | 721 | .924 | .918 | .014 | .106 | .590 | .210 | .03 |
| **b. Second random half** | 9,685 | 721 | .920 | .914 | .014 | .108 | .620 | .210 | .03 |
| **9.5.4.** Six correlated factors (AN on D; BN on D and E) | 17,880 | 723 | .924 | .918 | .014 | .1 | .614 | .168 | .02 |
| **a. First random half** | 9,293 | 723 | .924 | .918 | .014 | .107 | .620 | .160 | .02 |

| | | | | | | | | | |
|---|---|---|---|---|---|---|---|---|---|
| **b. Second random half** | 9,686 | 723 | .920 | .914 | .014 | .108 | .630 | .170 | .02 |
| **9.5.5.** Six correlated factors (AN on T; BN on E) | 18,299 | 723 | .922 | .916 | .014 | .11 | .615 | .166 | .02 |
| **a. First random half** | 9,559 | 723 | .922 | .916 | .014 | .109 | .610 | .160 | .02 |
| **b. Second random half** | 9,891 | 723 | .918 | .912 | .014 | .112 | .620 | .170 | .02 |
| **9.5.6.** Six correlated factors (OCD and OCPD on F) | 16,740 | 685 | .928 | .922 | .014 | .1 | .654 | .178 | .02 |
| **a. First random half** | 8,821 | 685 | .928 | .922 | .014 | .102 | .650 | .170 | .02 |
| **b. Second random half** | 8,988 | 685 | .926 | .920 | .014 | .101 | .640 | .180 | .02 |
| **9.5.7.** Six correlated factors (OCD and OCPD on N) | 16,765 | 685 | .928 | .922 | .014 | .1 | .611 | .182 | .02 |
| **a. First random half** | 8,842 | 685 | .928 | .922 | .014 | .101 | .640 | .180 | .02 |
| **b. Second random half** | 9,030 | 685 | .925 | .919 | .014 | .101 | .640 | .180 | .02 |
| **9.5.8.** Six correlated factors (OCD and OCPD on F and N) | 16,418 | 683 | .930 | .924 | .013 | .1 | .606 | .183 | .02 |
| **a. First random half** | 8,656 | 683 | .929 | .924 | .014 | .100 | .600 | .180 | .02 |
| **b. Second random half** | 8,858 | 683 | .927 | .921 | .014 | .100 | .620 | .190 | .02 |
| **9.5.9.** Six correlated factors (OCD and OCPD on D) | 16,739 | 685 | .928 | .922 | .014 | .1 | .637 | .177 | .02 |
| **a. First random half** | 8,830 | 685 | .928 | .922 | .014 | .101 | .640 | .170 | .02 |
| **b. Second random half** | 8,972 | 685 | .926 | .920 | .014 | .101 | .640 | .180 | .02 |
| **9.5.10.** Six correlated factors (BPD on E) | 17,244 | 685 | .926 | .920 | .014 | .1 | .611 | .176 | .02 |
| **a. First random half** | 9,062 | 685 | .926 | .920 | .014 | .106 | .600 | .170 | .02 |
| **b. Second random half** | 9,210 | 685 | .924 | .917 | .014 | .106 | .610 | .180 | .02 |
| **9.5.11.** Six correlated factors (BPD on D) | 17,775 | 685 | .924 | .917 | .014 | .1 | .619 | .179 | .02 |
| **a. First random half** | 9,367 | 685 | .923 | .917 | .014 | .107 | .640 | .170 | .02 |
| **b. Second random half** | 9,478 | 685 | .921 | .915 | .014 | .107 | .640 | .180 | .02 |
| **9.5.12.** Six correlated factors (BPD on D and E) | 17,051 | 684 | .927 | .921 | .014 | .1 | .617 | .172 | .02 |
| **a. First random half** | 8,974 | 684 | .927 | .921 | .014 | .106 | .600 | .170 | .02 |
| **b. Second random half** | 9,120 | 684 | .924 | .918 | .014 | .106 | .620 | .170 | .02 |
| **9.5.13.** Six correlated factors (BPD on D and T) | 16,773 | 684 | .928 | .922 | .014 | .1 | .615 | .177 | .02 |
| **a. First random half** | 8,819 | 684 | .928 | .922 | .014 | .106 | .610 | .170 | .02 |
| **b. Second random half** | 9,022 | 684 | .925 | .919 | .014 | .106 | .630 | .180 | .02 |
| **9.5.14.** Six correlated factors (BPD on E and T) | 17,238 | 684 | .926 | .920 | .014 | .1 | .61 | .188 | .02 |
| **a. First random half** | 9,056 | 684 | .926 | .920 | .014 | .106 | .600 | .180 | .02 |
| **b. Second random half** | 9,210 | 684 | .924 | .917 | .014 | .106 | .610 | .200 | .02 |
| **9.5.15.** Six correlated factors (Tic and TS on D) | 18,548 | 685 | .920 | .914 | .014 | .11 | .619 | .185 | .02 |
| **a. First random half** | 9,615 | 685 | .921 | .915 | .014 | .108 | .640 | .170 | .02 |
| **b. Second random half** | 9,916 | 685 | .917 | .911 | .015 | .109 | .630 | .180 | .02 |
| **9.5.16.** Six correlated factors (Tic and TS on N and D) | 18,275 | 683 | .921 | .915 | .014 | .1 | .634 | .237 | .02 |
| **a. First random half** | 9,565 | 683 | .921 | .915 | .014 | .107 | .620 | .200 | .03 |
| **b. Second random half** | 9,774 | 683 | .919 | .912 | .014 | .107 | .630 | .230 | .03 |
| **9.5.17.** Six correlated factors (ASPD on E and S) | 17,904 | 684 | .923 | .917 | .014 | .1 | .614 | .172 | .02 |
| **a. First random half** | 9,392 | 684 | .923 | .917 | .014 | .106 | .600 | .160 | .02 |
| **b. Second random half** | 9,602 | 684 | .920 | .914 | .014 | .106 | .630 | .170 | .02 |
| **9.5.26.** Six correlated factors (SCZ ~~ SCZAFF; OCD ~~ OCPD; MDD ~~ Dysthymia) | 15,962 | 682 | .932 | .926 | .013 | .1 | .617 | .174 | .02 |
| **a. First random half** | 8,431 | 682 | .931 | .926 | .013 | .102 | .630 | .170 | .02 |
| **b. Second random half** | 8,588 | 682 | .929 | .923 | .013 | .103 | .630 | .170 | .02 |
| **9.5.26.1.** Six correlated factors (PTSD on T) | 14,537 | 682 | .938 | .933 | .013 | .1 | .616 | .169 | .02 |
| **a. First random half** | 7,882 | 682 | .936 | .931 | .013 | .102 | .620 | .160 | .02 |
| **b. Second random half** | 7,717 | 682 | .937 | .932 | .013 | .102 | .630 | .170 | .02 |
| **9.5.26.2.** Six correlated factors (PTSD on T and D) | 13,428 | 681 | .943 | .938 | .012 | .1 | .614 | .175 | .02 |
| **a. First random half** | 7,203 | 681 | .942 | .937 | .012 | .102 | .610 | .170 | .02 |
| **b. Second random half** | 7,277 | 681 | .941 | .936 | .012 | .102 | .620 | .180 | .02 |
| **9.5.26.2.1.** Six correlated factors (OCD on T and D) | 12,147 | 680 | .949 | .944 | .011 | .09 | .611 | .188 | .02 |
| **a. First random half** | 6,609 | 680 | .948 | .943 | .012 | .100 | .600 | .180 | .02 |
| **b. Second random half** | 6,595 | 680 | .947 | .942 | .012 | .099 | .630 | .190 | .02 |
| **9.5.26.2.1.1** Six correlated factors (BPD on T and D) | 10,858 | 679 | .954 | .950 | .011 | .09 | .605 | .185 | .02 |
| **a. First random half** | 5,963 | 679 | .953 | .949 | .011 | .098 | .600 | .180 | .02 |
| **b. Second random half** | 5,957 | 679 | .953 | .948 | .011 | .098 | .610 | .190 | .02 |
| **9.5.26.2.1.1.1.** Six correlated factors (ED on T and D) | 9,469 | 678 | .961 | .957 | .01 | .09 | .598 | .202 | .02 |
| **a. First random half** | 5,261 | 678 | .959 | .956 | .010 | .097 | .600 | .190 | .02 |
| **b. Second random half** | 5,265 | 678 | .959 | .955 | .010 | .096 | .590 | .200 | .02 |

*Note.* CFI – comparative fit index, TLI – tucker-lewis index, RMSEA - root mean square error of approximation, SRMR – standardized root mean squared residual, FL – median factor loading, SD – standard deviation of loadings, SE – mean standard error. SCZ – schizophrenia, SCZAFF – schizoaffective, CD – conduct disorder, ODD – oppositional defiant disorder, PANIC – panic disorder, AGOR – agoraphobia. First half N = 63,981, second half N = 63,982.

**eTable 9**. *Split-half replication of exploratory bifactor models*

| Model | $\chi^2$ | df | CFI | TLI | RMSEA | SRMR | Alternative Indices FL | SD | SE |
|---|---|---|---|---|---|---|---|---|---|
| **2.** I, E, T | 12160.766 | 558 | 0.947 | 0.94 | 0.013 | 0.097 | 0.3 | 0.37 | 0.02 |
| **a. First Random Half** | 4254.175 | 552 | 0.966 | 0.962 | 0.01 | 0.088 | 0.59 | 0.18 | |
| **b. Second Random Half** | 3862.891 | 552 | 0.97 | 0.965 | 0.01 | 0.085 | 0.35 | 0.41 | |
| **3.** I, E, T, S | 12168.579 | 558 | 0.947 | 0.94 | 0.013 | 0.094 | 0.43 | 0.31 | 0.02 |
| **a. First Random Half** | 3170.741 | 548 | 0.976 | 0.973 | 0.009 | 0.081 | 0.39 | 0.42 | |
| **b. Second Random Half** | 3327.887 | 548 | 0.975 | 0.971 | 0.009 | 0.081 | 0.38 | 0.37 | |
| **4.** I, E, T, N | 14789.321 | 663 | 0.937 | 0.929 | 0.013 | 0.098 | 0.36 | 0.34 | 0.02 |
| **a. First Random Half** | 4614.839 | 653 | 0.965 | 0.96 | 0.01 | 0.088 | 0.53 | 0.18 | |
| **b. Second Random Half** | 4196.918 | 653 | 0.968 | 0.964 | 0.009 | 0.083 | 0.35 | 0.41 | |
| **5.** I, E, T, S, N | 15459.462 | 663 | 0.934 | 0.926 | 0.013 | 0.099 | 0.46 | 0.31 | 0.03 |
| **a. First Random Half** | 3578.646 | 648 | 0.974 | 0.97 | 0.008 | 0.082 | 0.39 | 0.49 | |
| **b. Second Random Half** | 3705.628 | 648 | 0.973 | 0.969 | 0.009 | 0.082 | 0.42 | 0.38 | |
| **6.** F, D, E, T | 17248.785 | 558 | 0.924 | 0.914 | 0.015 | 0.101 | 0.3 | 0.36 | 0.02 |
| **a. First Random Half** | 3871.018 | 548 | 0.97 | 0.965 | 0.01 | 0.087 | 0.34 | 0.49 | |
| **b. Second Random Half** | 3811.332 | 548 | 0.97 | 0.966 | 0.01 | 0.085 | 0.34 | 0.41 | |
| **7.** F, D, E, T, S | 16993.299 | 558 | 0.925 | 0.915 | 0.015 | 0.099 | 0.39 | 0.32 | 0.02 |
| **a. First Random Half** | 3138.617 | 543 | 0.977 | 0.973 | 0.009 | 0.081 | 0.38 | 0.41 | |
| **b. Second Random Half** | 3279.643 | 543 | 0.975 | 0.971 | 0.009 | 0.081 | 0.38 | 0.36 | |
| **8.** F, D, E, T, N | 19485.974 | 661 | 0.916 | 0.906 | 0.015 | 0.1 | 0.36 | 0.32 | 0.03 |
| **a. First Random Half** | 4172.646 | 646 | 0.969 | 0.964 | 0.009 | 0.086 | 0.32 | 0.56 | |
| **b. Second Random Half** | 4144.169 | 646 | 0.969 | 0.964 | 0.009 | 0.083 | 0.39 | 0.47 | |
| **9.** F, D, E, T, S, N | 20171.114 | 663 | 0.913 | 0.903 | 0.015 | 0.102 | 0.42 | 0.3 | 0.03 |
| **a. First Random Half** | 3549.477 | 642 | 0.974 | 0.97 | 0.008 | 0.082 | 0.45 | 0.48 | |
| **b. Second Random Half** | 3683.517 | 642 | 0.973 | 0.969 | 0.009 | 0.08 | 0.55 | 0.87 | |
| **9.5.** Six correlated factors (Hyp-Imp on E; Inattentive and Combined on N) | 17354.233 | 661 | 0.925 | 0.916 | 0.014 | 0.097 | 0.4 | 0.29 | 0.03 |
| **a. First Random Half** | 3579.357 | 640 | 0.974 | 0.97 | 0.008 | 0.082 | 0.62 | 0.2 | |
| **b. Second Random Half** | 3462.609 | 640 | 0.975 | 0.971 | 0.008 | 0.078 | 0.6 | 0.22 | |
| **9.5.26.** Six correlated factors (SCZ ~~ SCZAFF; OCD ~~ OCPD; MDD ~~ Dysthymia) | 17068.688 | 658 | 0.927 | 0.917 | 0.014 | 0.095 | 0.42 | 0.27 | 0.03 |
| **a. First random half** | 3133.882 | 637 | 0.978 | 0.974 | 0.008 | 0.081 | 0.62 | 0.2 | |
| **b. Second random half** | 2982.571 | 637 | 0.979 | 0.976 | 0.008 | 0.077 | 0.58 | 0.22 | |
| **9.5.26.2.1.1.1.** Six correlated factors (ED on T and D) | 16359.642 | 654 | 0.93 | 0.92 | 0.014 | 0.095 | 0.38 | 0.28 | 0.03 |
| **a. First random half** | 2991.875 | 633 | 0.979 | 0.976 | 0.008 | 0.079 | 0.52 | 0.22 | |
| **b. Second random half** | 2915.415 | 633 | 0.98 | 0.976 | 0.008 | 0.076 | 0.55 | 0.24 | |

*Note*. CFI – comparative fit index, TLI – tucker-lewis index, RMSEA - root mean square error of approximation, SRMR – standardized root mean squared residual, FL – median factor loading, SD – standard deviation of loadings, SE – mean standard error. SCZ – schizophrenia, SCZAFF – schizoaffective, CD – conduct disorder, ODD – oppositional defiant disorder, PANIC – panic disorder, AGOR – agoraphobia. First half N = 63,981, second half N = 63,982.

**eTable 10**. *Split-half replication of exploratory modified bifactor models*

| | | | | | | | Alternative Indices | | |
|---|---|---|---|---|---|---|---|---|---|
| **Model** | **$\chi^2$** | **df** | **CFI** | **TLI** | **RMSEA** | **SRMR** | **FL** | **SD** | **SE** |
| **2.** I, E, T | 7004.995 | 555 | .971 | .967 | .01 | .081 | .31 | 0.37 | 0.02 |
| **a. First Random Half** | 3905.403 | 555 | .97 | .966 | .01 | .088 | .28 | 0.37 | 0.03 |
| **b. Second Random Half** | 3862.891 | 555 | .97 | .966 | .01 | .085 | .33 | 0.36 | 0.03 |
| **3.** I, E, T, S | 5756.216 | 552 | .976 | .973 | .009 | .076 | .38 | 0.33 | 0.02 |
| **a. First Random Half** | 3170.741 | 552 | .976 | .973 | .009 | .081 | .36 | 0.33 | 0.03 |
| **b. Second Random Half** | 3327.887 | 552 | .975 | .971 | .009 | .081 | .38 | 0.34 | 0.03 |
| **4.** I, E, T, N | 7466.448 | 657 | .97 | .966 | .009 | .08 | .35 | 0.35 | 0.02 |
| **a. First Random Half** | 4614.839 | 657 | .965 | .961 | .01 | .088 | .51 | 0.18 | 0.03 |
| **b. Second Random Half** | 4196.918 | 657 | .968 | .964 | .009 | .083 | .35 | 0.35 | 0.03 |
| **5.** I, E, T, S, N | 6350.994 | 653 | .975 | .971 | .008 | .077 | .43 | 0.33 | 0.02 |
| **a. First Random Half** | 3842.144 | 653 | .972 | .968 | .009 | .085 | .59 | 0.15 | 0.03 |
| **b. Second Random Half** | 3705.628 | 653 | .973 | .969 | .009 | .082 | .41 | 0.33 | 0.03 |
| **6.** F, D, E, T | 6922.238 | 552 | .971 | .967 | .009 | .08 | .32 | 0.37 | 0.02 |
| **a. First Random Half** | 3871.018 | 552 | .97 | .966 | .01 | .087 | .29 | 0.38 | 0.03 |
| **b. Second Random Half** | 3811.332 | 552 | .97 | .966 | .01 | .085 | .33 | 0.37 | 0.03 |
| **7.** F, D, E, T, S | 5680.54 | 548 | .977 | .973 | .009 | .075 | .39 | 0.34 | 0.02 |
| **a. First Random Half** | 3138.617 | 548 | .977 | .973 | .009 | .081 | .36 | 0.34 | 0.03 |
| **b. Second Random Half** | 3279.643 | 548 | .975 | .971 | .009 | .081 | .38 | 0.34 | 0.03 |
| **8.** F, D, E, T, N | 7374.768 | 651 | .97 | .966 | .009 | .08 | 0.32 | 0.36 | 0.02 |
| **a. First Random Half** | 4568.608 | 651 | .965 | .961 | .01 | .087 | 0.5 | 0.22 | 0.03 |
| **b. Second Random Half** | 4394.246 | 651 | .966 | .962 | .009 | .082 | 0.51 | 0.26 | 0.03 |
| **9.** F, D, E, T, S, N | 6285.615 | 648 | .975 | .971 | .008 | .077 | 0.42 | 0.33 | 0.02 |
| **a. First Random Half** | 3795.468 | 648 | .972 | .968 | .009 | .085 | 0.61 | 0.15 | 0.03 |
| **b. Second Random Half** | 3683.517 | 648 | .973 | .969 | .009 | .08 | 0.57 | 0.16 | 0.03 |
| **9.5.** Six correlated factors (Hyp-Imp on E; Inattentive and Combined on N) | 5766.904 | 646 | .977 | .974 | .008 | .072 | 0.43 | 0.31 | 0.02 |
| **a. First Random Half** | 3579.357 | 646 | .974 | .97 | .008 | .082 | 0.58 | 0.19 | 0.03 |
| **b. Second Random Half** | 3462.609 | 646 | .975 | .971 | .008 | .078 | 0.6 | 0.21 | 0.03 |
| **9.5.26.** Six correlated factors (SCZ ~~ SCZAFF; OCD ~~ OCPD; MDD ~~ Dysthymia) | 5094.605 | 643 | .98 | .977 | .007 | .07 | 0.4 | 0.3 | 0.02 |
| **a. First random half** | 3133.882 | 643 | .978 | .975 | .008 | .081 | 0.58 | 0.18 | 0.03 |
| **b. Second random half** | 2982.571 | 643 | .979 | .976 | .008 | .077 | 0.58 | 0.2 | 0.03 |
| **9.5.26.2.1.1.1.** Six correlated factors (ED on T and D) | 4912.169 | 639 | .981 | .978 | .007 | .069 | 0.4 | 0.51 | 0.08 |
| **a. First random half** | 2991.875 | 639 | .979 | .976 | .008 | .079 | 0.5 | 0.2 | 0.03 |
| **b. Second random half** | 2915.415 | 639 | .98 | .976 | .007 | .076 | 0.5 | 0.22 | 0.03 |

*Note*. CFI – comparative fit index, TLI – tucker-lewis index, RMSEA - root mean square error of approximation, SRMR – standardized root mean squared residual, FL – median factor loading, SD – standard deviation of loadings, SE – mean standard error. SCZ – schizophrenia, SCZAFF – schizoaffective, CD – conduct disorder, ODD – oppositional defiant disorder, PANIC – panic disorder, AGOR – agoraphobia. First half N = 63,981, second half N = 63,982.

**eTable 11.** *External correlates of the higher-order psychopathology dimensions*

| | Externalizing Beta [95% CI] | Substance Use Beta [95% CI] | Thought Problems Beta [95% CI] | Fear Beta [95% CI] | Distress Beta [95% CI] | NDD Beta [95% CI] |
|---|---|---|---|---|---|---|
| **Sex** | .204 [.153 - .255] | .559 [.540 - .578] | .079 [.057 - .102] | -.348 [-.379 - -.317] | -.390 [-.408 - -.372] | .056 [.022 - .090] |
| **Sleep Disorder** | .300 [.232 - .368] | -.067 [-.102 - -.033] | .148 [.113 - .183] | .415 [.374 - .455] | .611 [.583 - .639] | .379 [.332 - .427] |
| **Income** | -.081 [-.093 - -.069] | -.257 [-.262 - -.252] | -.200 [-.206 - -.194] | -.062 [-.068 - -.055] | -.067 [-.071 - -.063] | .003 [-.005 - .011] |
| **Education** | -.129 [-.155 - -.103] | -.423 [-.433 - -.413] | -.237 [-.249 - -.226] | -.052 [-.067 - -.038] | -.040 [-.049 - -.031] | .100 [.081 - .108] |

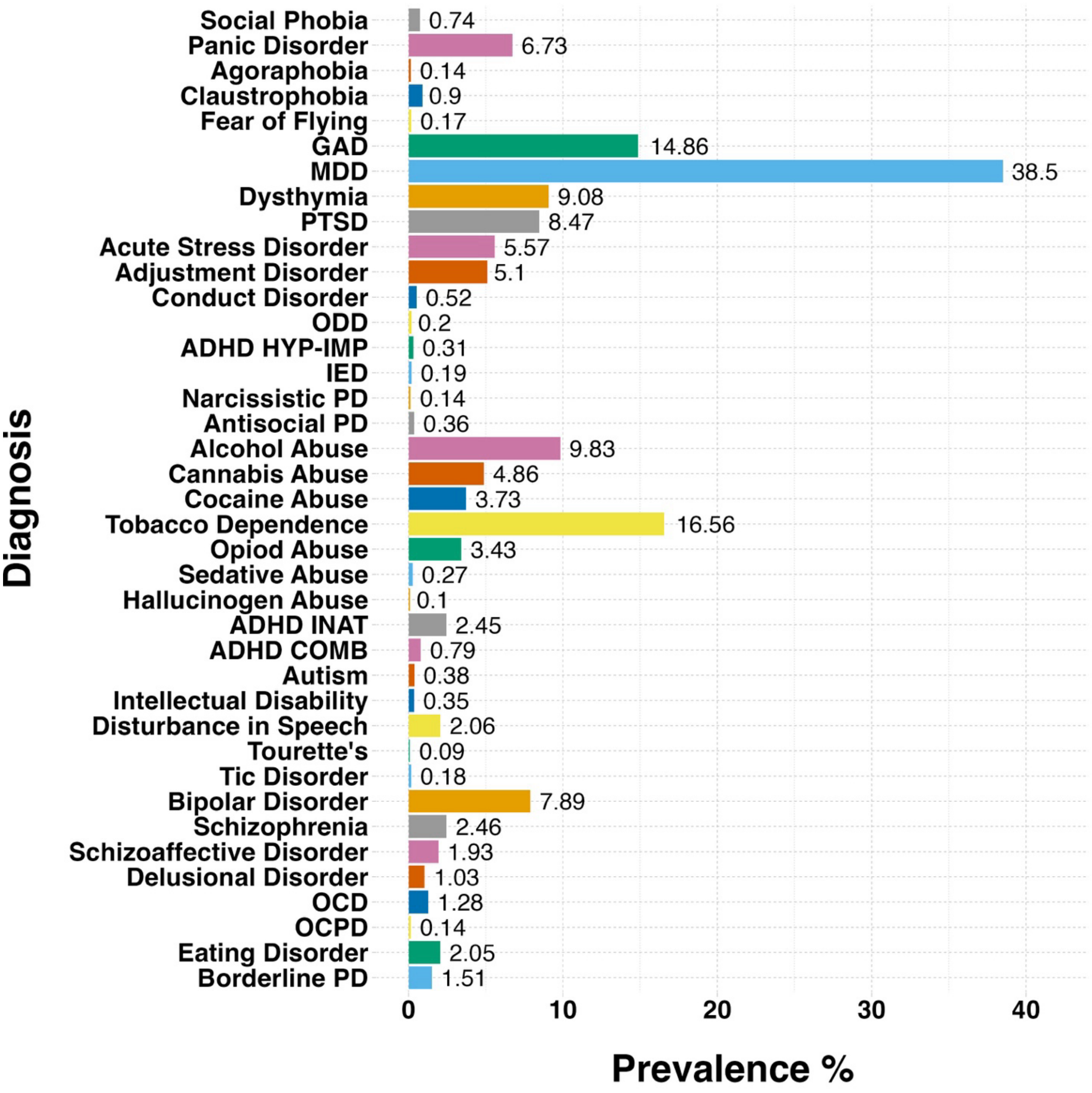


**eFigure 1**. *Prevalences of the 39 primary lifetime diagnoses*.

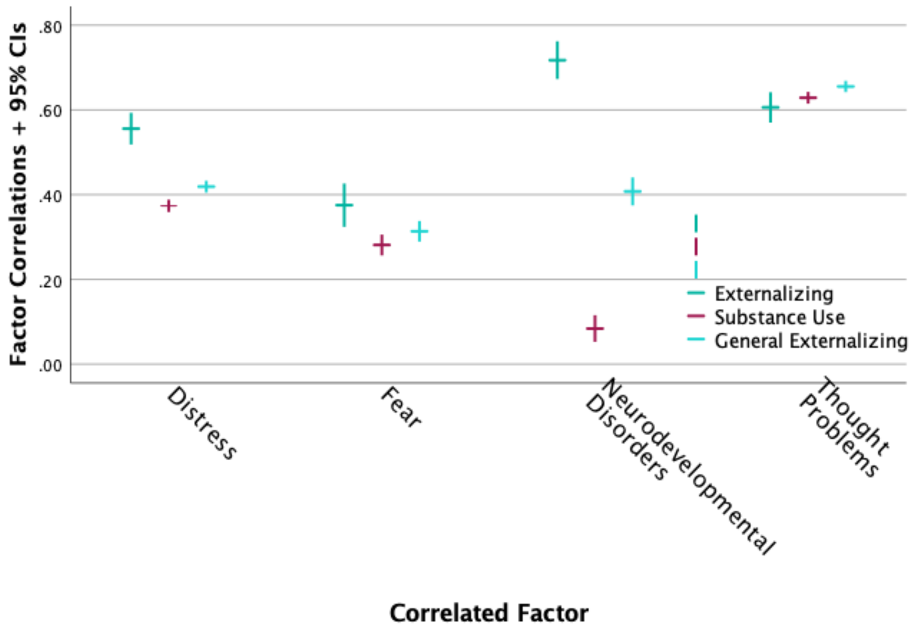


**eFigure 2a.** *Correlations of D, F, N, and T factors with Distinct Externalizing, Substance Use, and Combined Externalizing + Substance Use factors (General Externalizing)*

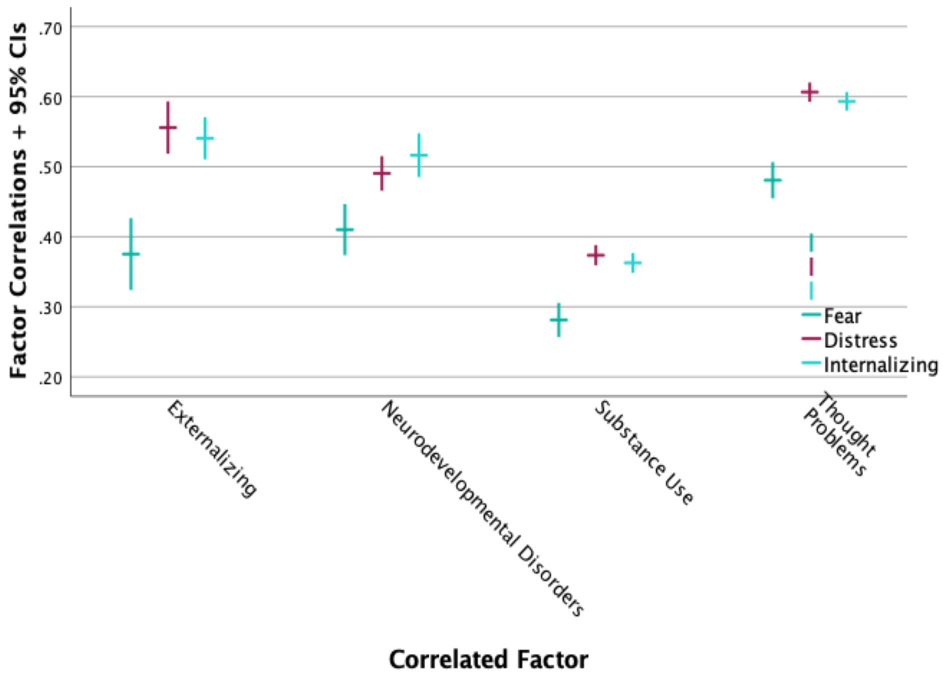


**eFigure 2b.** *Correlations of E, N, S, and T factors with Fear, Distress, and a broad Internalizing factor*

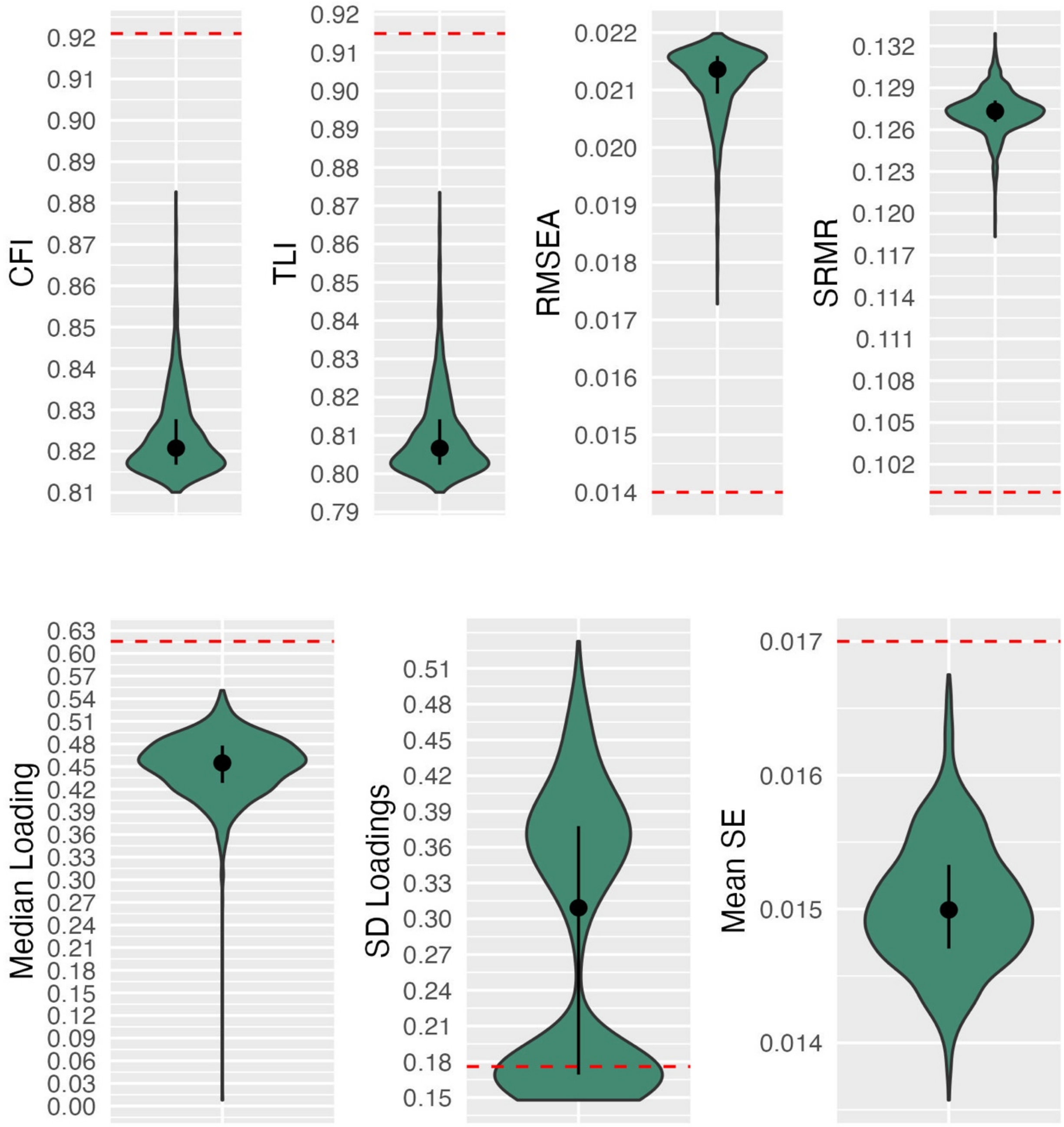


**eFigure 3.** *Indices of robustness of the results of the best-fitting model using random models***.**
*Note*. Red dashed line represents the fit of the best fitting CFA model (Model 9.5 with ASPD on S). The violins represent the distribution of the indices from the 1,000 random models.
